# Disaggregating proportional multistate lifetables by population heterogeneity to estimate intervention impacts on inequalities

**DOI:** 10.1101/2021.01.24.21250411

**Authors:** Patrick Andersen, Anja Mizdrak, Nick Wilson, Anna Davies, Laxman Bablani, Tony Blakely

## Abstract

**Background:** Simulation models can be used to quantify the projected health impact of interventions. Quantifying heterogeneity in these impacts, for example by socioeconomic status, is important to understand impacts on health inequalities.

We aim to disaggregate one type of Markov macro-simulation model, the proportional multistate lifetable, ensuring that under business-as-usual (BAU) the sum of deaths across disaggregated strata in each time step returns the same as the initial non-disaggregated model. We then demonstrate the application by deprivation quintiles for New Zealand (NZ), for: hypothetical interventions (50% lower all-cause mortality, 50% lower coronary heart disease mortality) and a dietary intervention to substitute 59% of sodium with potassium chloride in the food supply.

**Methods:** We developed a disaggregation algorithm that iteratively rescales mortality, incidence and case fatality rates by time-step of the model to ensure correct total population counts were retained at each step.

To demonstrate the algorithm on deprivation quintiles in NZ, we used the following inputs: overall (non-disaggregated) all-cause mortality &morbidity rates, coronary heart disease incidence &case fatality rates; stroke incidence &case fatality rates. We also obtained rate ratios by deprivation for these same measures. Given all-cause and cause-specific mortality rates by deprivation quintile, we derived values for the incidence, case fatality and mortality rates for each quintile, ensuring rate ratios across quintiles and the total population mortality and morbidity rates were returned when averaged across groups.

The three interventions were then run on top of these scaled BAU scenarios.

**Results:** The algorithm exactly disaggregated populations by strata in BAU. The intervention scenario life years and health adjusted life years (HALYs) gained differed slightly when summed over the deprivation quintile compared to the aggregated model, due to the stratified model (appropriately) allowing for differential background mortality rates by strata. Modest differences in health gains (health adjusted life years) resulted from rescaling of sub-population mortality and incidence rates to ensure consistency with the aggregate population.

**Conclusion:** Policy makers ideally need to know the effect of population interventions estimated both overall, and by socioeconomic and other strata. We demonstrate a method and provide code to do this routinely within proportional multistate lifetable simulation models and similar Markov models.

## Background

Quantifying the health impacts of population interventions by social strata is necessary for the design of effective policies to reduce health inequalities. These analyses are often based on simulation models, which attempt to quantify the future health (and cost) impacts of interventions. For example: what is the health gain of various tobacco (1), diet (2–5) and other preventive interventions by socioeconomic groups? And, how will the intervention change health inequalities?

Simulating interventions by socioeconomic group is difficult. The heterogeneity of epidemiological parameters (e.g. incidence, mortality, morbidity, case fatality, etc.) across socioeconomic strata and determining their effects on simulation outputs is challenging; both in specifying the relevant inputs correctly, and coherence in the modeling (e.g. ensuring that deaths and other outputs such as prevalence sum across strata to return that in the aggregated population).

For an illustrative example, consider a lifetable simulation for a population of 2000 members. Suppose that the mortality risk of the population is 1 per 100 and increases by 20% per year for 20 years to account for aging. After the first year, there will be an expected 1980 people alive, and by the 20^th^ year, the expected population will decrease to 367.4. The person-years lived by annual cycle are shown in Figure 1.

**Figure 1.**
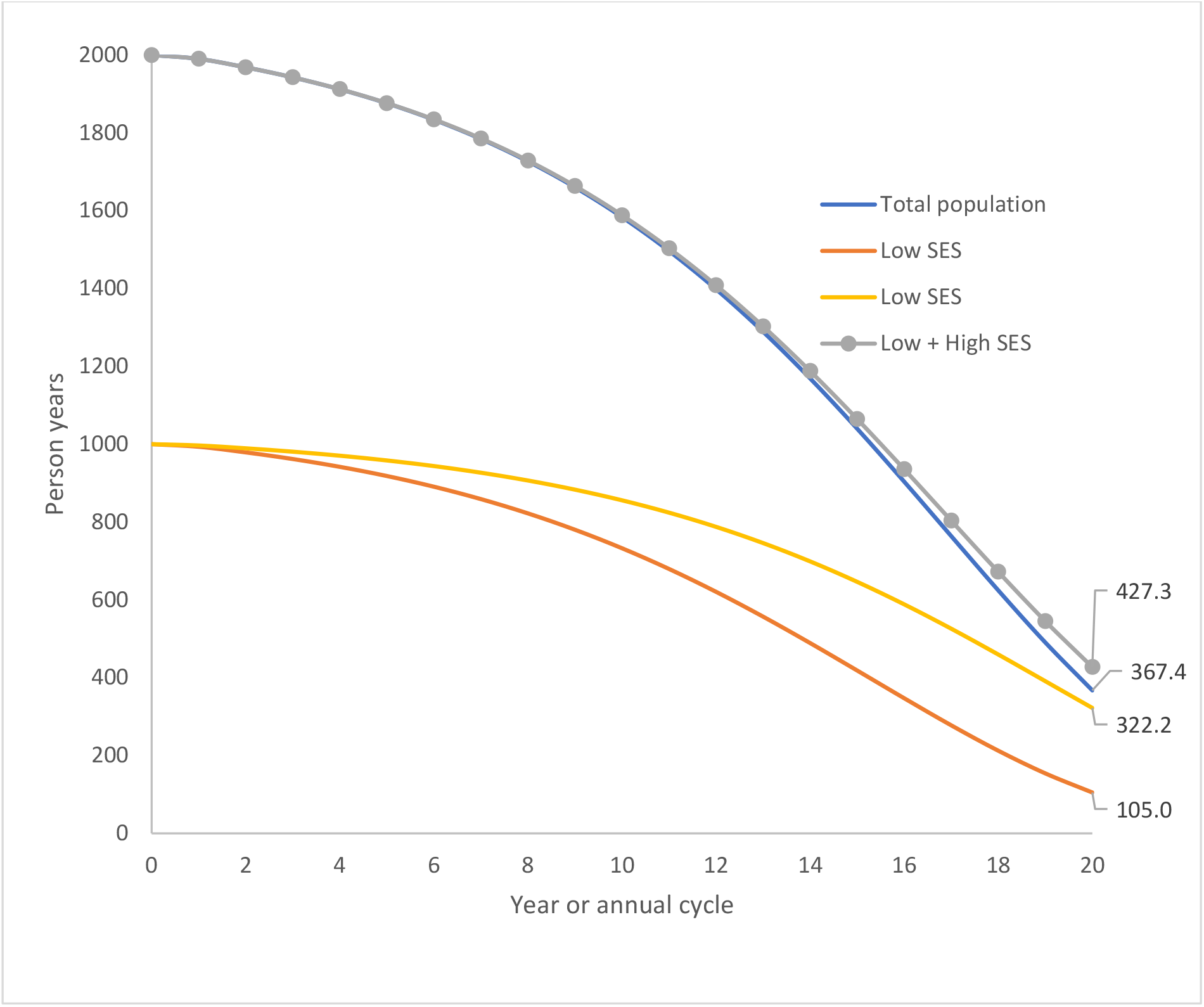
Demonstration of the principle issues addressed in our study: Persons years lived by annual cycle for a population of initial size n=2000, with mortality rate 1 per 100 person years in the first year, and the high and low SES groups (each N=1000) that make up this total population. The mortality rate in the low SES group is twice that in the high SES group at baseline (i.e. 0.667 and 1.333 per 100 person years, respectively). The mortality rate increase by 20% per annum in the total population, and each SES group.

Now suppose that our population is comprised of two distinct groups, e.g., high and low socioeconomic status (SES) groups, each with 1000 people. Suppose also that one group has twice the mortality risk of the other. Setting the mortality risks for the groups to be 4/3 and 2/3 deaths per 100 respectively yields the same expected number of living people after the first year as were obtained previously (i.e., 1980). By applying the annual 20% increase of mortality risk to each group and simulating to the end of 20^th^ year, we generate the number of person years as shown in Figure 1. In the 20^th^ annual cycle there are 322.2 and 105.0 expected person years lived in the high and low mortality group respectively. This sums to 427.3, which is greater than the 367.4 obtained when the population was modeled as a whole. That is, the mortality parameters are mis-specified in the stratified model, because differing mortality rates by strata change the proportional distribution of people across strata away from 50:50 over time. An intervention scenario imposed on top of these mis-specified lifetables likewise incorrectly estimate the health gains for each stratum and the differences in health gain by stratum (i.e. the inequality impact).

The purpose of our study was to develop methods to disaggregate populations in Markov macro-simulation models, particularly proportional multistate lifetable models (PMSLT), (6,7), retaining fidelity with the aggregate population in terms of numbers of deaths and other event counts. We searched the literature for previous publications on this topic but could find none (see Appendix 1).

A few extensions of multistate models accounting for heterogeneity have been observed in the literature. Such extensions have highlighted, in a disease-free context with independent populations, that mortality is underestimated if heterogeneity is not accounted for (8,9). Further, log-linear, hazard, or Bayesian regression (10) methods can be used to calculate differences in multistate life table parameter values by an explicitly defined heterogenous parameter of interest (say, SES or education) for further simulation. Other related works in epidemiology and demography demonstrate methods for obtaining the mortality of diseased and non-diseased cohorts from the overall mortality rate in continuous time MSLT models (11,12).

Our work differs from the previous literature in that we use a proportional version of multistate lifetables where rates only vary at discrete time steps. We also treat the aggregate population as the ‘working truth’, with our task being to obtain a unique set of valid rates for the strata in order to quantify the impacts on health inequalities, e.g. socioeconomic position.

First, we outline the mathematics of disaggregating populations, such that separately simulating the transitions of each sub-stratum under BAU through its state transition Markov chain yields death counts and person-years lived that are identical with the aggregate population. Second, we demonstrate intervention scenarios applied to both the aggregate and deprivation-stratified models. We deliberately chose interventions that do not have differing effect sizes by deprivation quintile. The three modeled scenarios are two hypothetical interventions (50% lower all-cause mortality, 50% lower coronary heart disease mortality) and a dietary intervention to substitute 59% of sodium chloride with potassium chloride in the food supply in New Zealand.

## Methods

We consider two separate Markov processes which describe the mortality and disease lifetables used in PMSLT modeling (6,7).

### 1.1 The Mortality Lifetable

The *mortality lifetable* of a population *P* measures metrics such as the number of deaths, life years and life expectancy, capturing snapshots at discrete timesteps (e.g. years). Let *A*_*t*_ and *D*_*t*_ denote the number of people alive and dead at the end of the *t*-th timestep. We assume *A*_0_ = *N* and *D*_0_ = 0, where *N* is the total number of people that are observed initially, and for each *t* we have a mortality rate *m*_*t*_. Then:

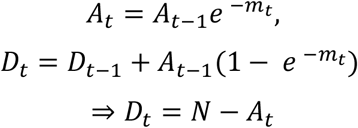

In the *mortality lifetable disaggregation problem, P* consists of *n* underlying sub-populations *P*_1_, …, *P*_*n*_, where we assume that members of *P* remain in their respective sub-populations for life. The initial population for each *P*_1_, …, *P*_*n*_ are known, and the initial mortality rates are known (either given, or solvable using mortality rate ratios between strata). However, whilst *m*_*t*_, the total population mortality rate by future time step, is given, the stratum-specific mortality rates over time are not given. This situation is not uncommon, e.g. we may know the starting disaggregation of a population by socioeconomic strata, and the rate ratios for mortality or disease incidence comparing strata, but not the exact rates by strata over time. The goal is to use the aggregate populations mortality rates, the rate ratios comparing strata, and starting distribution of each sub-population (*P*_*k*_) to solve the mortality lifetables for each *P*_1_, …, *P*_*n*_. The sub-populations lifetables must be *consistent* with that of the aggregate population: at each time timestep *t*, the sum of the people in the alive (dead) compartment for each sub-population is equal to the number of people in the alive (dead) compartment for the aggregate population. i.e.,

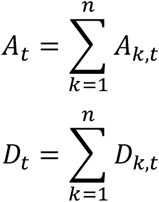

where *A*_*k,t*_ and *D*_*k,t*_ denote the number of people in the alive and dead compartments respectively for sub-population *k* at time *t*. Thus, the disaggregation problem reduces to finding mortality rates *m*_*k,t*_ for each sup-population *k* at timestep *t* > 0 such that:

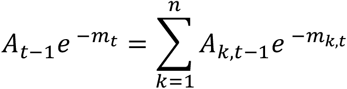

If we let *P*_1_ be the reference sub-population, then the mortality rate ratios *r* for timestep *t* are given as scalars 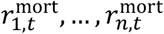, where 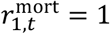, such that 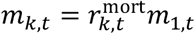 for each sub-population *k*. By substituting each 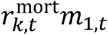 into the consistency equations, we obtain a set of equations with a unique solution. By solving these, we then able to obtain a unique set of sub-population mortality rates for the mortality lifetable disaggregation problem. A method for solving for these rates, and the proof of the uniqueness of the solution is in Appendix 2.

### 1.2 The Mortality/Morbidity Lifetable

We now extend the mortality lifetable to a *mortality/morbidity lifetable* that also includes HALYs which incorporate the effects of morbidity. Let *L*_*t*_ denote population life years at *t*, where:

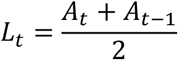

Our HALY unit is a rescaling of the life-year using disability rates. Let *w*_*t*_ denote the prevalent years lost to disability (pYLD) from a burden of disease study at time *t*, divided by the population in that strata. Then the formula for HALYs at time *t*, denoted 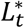, is:

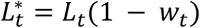

For the *mortality/morbidity lifetable disaggregation problem*, an extension of the problem in Section 1.1, we are given a mortality/morbidity lifetable for a population *P* which includes all parameters from the mortality lifetable, along with morbidity weights *w*_*t*_ and HALYs 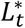. As before, *P* consists of *n* sub-populations *P*_1_, …, *P*_*n*_, with their individual population counts, mortality rates, and pYLDs given for the first time step. The goal is to use the aggregate lifetable to determine the mortality rates and pYLD rates for each sub-population *P*_*k*_, and hence obtain the mortality/morbidity lifetables for each *P*_1_, …, *P*_*n*_. We have to ensure alive population counts for sub-populations and the aggregate population agree and also total HALYs for the sub-populations agree with the aggregate population HALYs. That is:

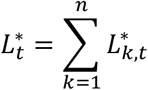

where 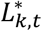 denotes the HALYs for sub-population *k* at time *t*. To satisfy the above equations, we must solve values *w*_*k,t*_ *for each k and t such* that:

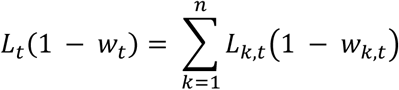

where *L*_*k,t*_ denotes life years for sub-population *k* at *t*.

We can assume that the mortality lifetable disaggregation problem has been solved as a subproblem, since it can be independently solved using the method in section 2.1. Then, we have alive population values, such that 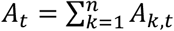, which implies that 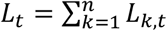. Hence, we can simplify the HALY constraints to:

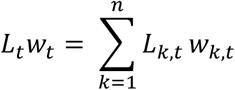

To solve the problem, we assume *pYLD ratios* for each *t* (although in all likelihood ratios vary by age and sex, but are assumed constant over t), which are 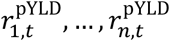, and 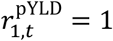, such that 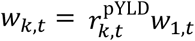 for each sub-population *k*. After substituting each 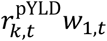 into the HALY constraints and solving, we obtain:

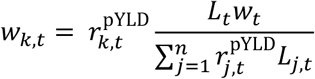

Thus, we are able to use pYLD ratios to uniquely disaggregate the mortality/morbidity lifetable such that the HALYs in the sub-population lifetables are consistent with the aggregate population lifetable.

### 1.3 The Disease Lifetable

A PMSLT, described in detail in (6,7), works through changes in disease incidence or case fatality rates, where each disease is assumed independent of other diseases. Similar to the all-cause mortality and morbidity lifetable (above), the issue here is in ensuring that each disease-specific subsidiary lifetable also returns the numbers and rates or the total population before it is disaggregated by heterogeneity (eg. SES).

The *disease lifetable* consists of three compartments: a healthy compartment *S*, diseased compartment *C*, and dead compartment *D*. At each timestep, members of the population in *S* transition to *C* according to the *incidence rate*, and from *C* to *D* according to the *fatality rate*. For some diseases, members can transition from *C* to *S* as per the *remission rate*, however, for simplicity, we do not consider this possibility for now.

Let *S*_*t*_, *C*_*t*_ and *D*_*t*_ denote the number of people in compartment *S, C* and *D* respectively at the end of the *t*-th step. We will assume initially that *D*_0_ = 0 and *S*_0_ + *C*_0_ = *N*, where *N* is the total number of people initially observed. Let *i*_*t*_ and *f*_*t*_ denote the incidence and fatality rates respectively at *t*. The equations for *S*_*t*_, *C*_*t*_ and *D*_*t*_ are given by the system:

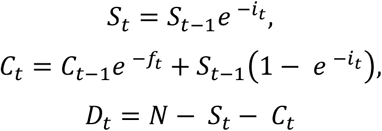

These equations are premised on a simplifying assumption that members of the population cannot die from the disease in the same timestep in which they contract the disease This assumption can, in practice, be mitigated through choosing an appropriately small timestep.

For the *disease lifetable disaggregation problem*, we are given the disease lifetable for an aggregate population *P* complete with incidence rates *i*_*t*_, fatality rates *f*_*t*_ and population counts *S*_*t*_, *C*_*t*_ and *D*_*t*_ at each timestep *t*. We assume that *P* consists of *n* separate underlying sub-populations *P*_1_, …, *P*_*n*_, each with their own population counts, incidence rates and fatality rates. Let *S*_*k,t*_, *C*_*k,t*_ and *D*_*k,t*_ denote the number of people in the healthy, diseased and dead compartments respectively for sub-population *k* at time *t*. We additionally assume for each sub-population we are given the initial disease prevalence, hence we can obtain *S*_*k*,0_ and *C*_*k*,0_. The objective of the problem is to determine both the incidence rates and fatality rates of each sub-population *P*_*k*_ and hence obtain the disease lifetables for each *P*_1_, …, *P*_*n*_. The criteria for consistency for this disaggregation at each time timestep *t* are given by:

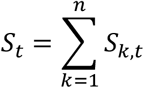

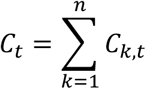

I.e., we must choose incidence rates *i*_*k,t*_ and fatality rates *f*_*k,t*_ for each timestep *t* and sub-population *k* such that:

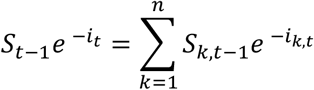

and

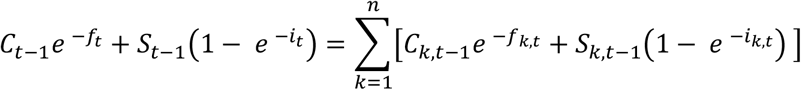

We once again assume that we are given rate ratios for the sub-population rates at each timestep *t*, specifically *incidence rate ratios* 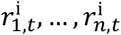 such that 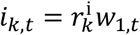 for each *k*, and *fatality rate ratios* 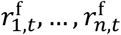 such that 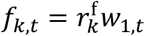 for each *k*.

We can apply the method used in the mortality problem to obtain unique incidence rates *i*_*k,t*_ that satisfy the constraints for the healthy population. After obtaining the sub-population incidence rates, the consistency constraint for the diseased population simplifies to:

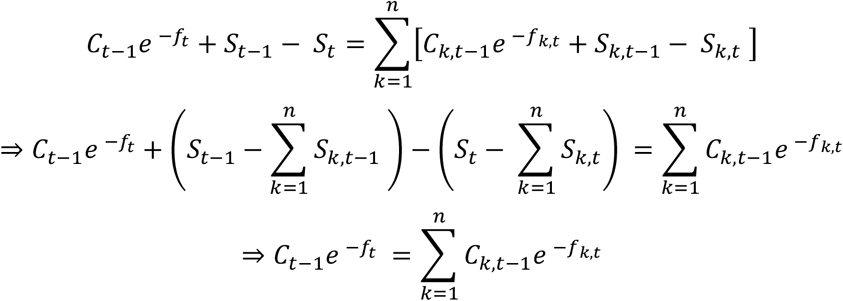

Thus, by using two consecutive applications of the methods described in Appendix 1, first for the healthy compartment and incidence rates and then for the diseased compartment and fatality rates, we can use the rate ratios to obtain a consistent disaggregation of the disease lifetable.

The prototype code for the above methods is provided in a GitHub repository (13).

### Case studies of interventions by deprivation strata

Our case studies are applied to the NZ population, which we disaggregate by small area deprivation (a census index at the geographic unit of about 100 individuals).

#### Intervention 1: 50% reduction in ACMR

Our first intervention is a hypothetical 50% reduction of the all-cause mortality rate (ACMR) for Māori females (Māori being the Indigenous population of NZ and suffering elevated levels of deprivation, and a determinant of SES hence we routinely stratify by sex, age and ethnicity prior to then stratifying by SES). This intervention acts directly upon the mortality/morbidity lifetable by modifying the ACMR and so does not require modeling of any disease lifetables. Supplementary Table 1 shows the aggregate ACMR and pYLD values for Māori females at each age and Supplementary Table 2 gives the aggregate population counts for each 5-year age group. We apply an annual percentage change in mortality (APC) of -2.5% to the ACMR values for each year after 2011 until 2026 (14).

To uniquely disaggregate the main lifetable for Māori females we use the rate ratios for five categories of deprivation obtained from routine health data for ACMR and pYLD values (Supplementary Tables 3 and 4 respectively). The initial proportions for the five strata are set at 20% each.

To apply the intervention, we multiply each business-as-usual (BAU) ACMR rate by 0.5 for both the aggregate and stratified populations.

### 1.4 Intervention 2: 50% reduction in CHD Incidence

In our second intervention, we reduce the incidence rate of coronary heart disease (CHD) for Māori females by 50%. This intervention requires the modeling of a CHD disease lifetable as well as the main mortality/morbidity lifetable. The incidence rate, fatality rate, disability rate and initial prevalence values for the aggregate population are given in Supplementary Table 5. We assume an APC of -2% for both incidence and fatality rates for each year after 2011 until 2026 (giving a 4% per annum reduction in mortality).

To disaggregate the CHD lifetable, we use the rate ratios for five categories of deprivation from routine health data for CHD incidence and fatality rates given in Supplementary Tables 6 and 7 respectively. We also use *prevalence ratios* for CHD given in Supplementary Table 8 to obtain initial prevalence values for the sub-populations.

To apply the intervention, we multiply the CHD incidence rate in BAU by 0.5 for both the aggregate and stratified populations. This change in incidence flows through into changes in CHD mortality and prevalence, and then to changes in all-cause mortality/morbidity in the main lifetable (as described in section 2.5).

### 1.5 Intervention 3: 59% Reduction in NZ sodium consumption

In our final example, we apply a real-world dietary intervention. As described in Nghiem et al. (15), we assume that 59% of the dietary sodium in processed foods and table salt is replaced by potassium and magnesium salts. This is estimated to reduce daily sodium intake by 51.5% for the NZ population. We assume that the effect size is the same across sub-populations (consistent with similar sodium intakes across the sub-populations considered here).

This intervention reduces the incidence rates of CHD and stroke (due to a reduction of systolic blood pressure). As such, our modeling involves both CHD and stroke disease lifetables. The CHD lifetables are obtained as per section 2.5. The stroke lifetables are similarly obtained using the aggregate values from Supplementary Table 9 and rate ratios for incidence, fatality and prevalence from Supplementary Tables 10, 11 and 12 respectively. We again assume an APC = -2% for the incidence and fatality rates of stroke for each year after 2011 until 2026.

The PSMLT modeling technique we use to apply this intervention to the disease lifetables uses *potential impact fractions (PIFs)* to scale the BAU disesase incidence rates. Our PIFs for each disease d ∈ {CHD, stroke}(7) are obtained using the RR shift method described in Barendregt and Veerman (16):

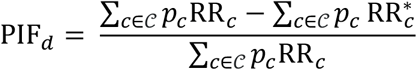

where 𝒞 is the set of risk exposure categories *c, p*_*c*_ is the population fraction in category *c*, and RR_*c*_ and 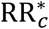 are the relative risks of *c* before and after the intervention respectively. The proportions of the female Māori population in sodium risk categories are obtained from our analysis of national nutrition survey data (17) and given in Supplementary Table 13. To determine RR_*c*_ and 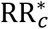, we apply the relative risks per 1g daily sodium increase, from Blakely et al. (5) (to the mean sodium intakes for each category) where the mean intakes for 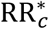 are obtained from a 51.5% reduction of the BAU mean intakes. Supporting tables are presented in Supplementary Table 14 and Supplementary Table 15.

## Results

Table 1 shows the PMSLT outputs in BAU for 60-64 year old Māori females (centered on age 62) alive in 2011, for the next 20 years until aged 82. Regarding ACMRs, there is no difference for the total population modeled in aggregate compared to the weighted average across deprivation heterogeneity strata – as there should be given the method described above. Similarly, the total HALYs by cycle and summed to age 80, are identical between the aggregate and disaggregated PMSLT. Also shown in Table 1 are the AMCR for the least and most deprived quintile (with a rate ratio of 1.5812 at age 62 decreasing to 1.1159 by age 82). Whilst the population distribution is 20% in each quintile of deprivation, the HALYs lived by the least deprived are greater than for the most deprived at all ages, and more so with increasing age such that summed from age 62 to 82 the least deprived have 121.9 (18.7%) more accrued HALYs than the most deprived – due to both higher morbidity and higher ACMR.

**Table 1:**
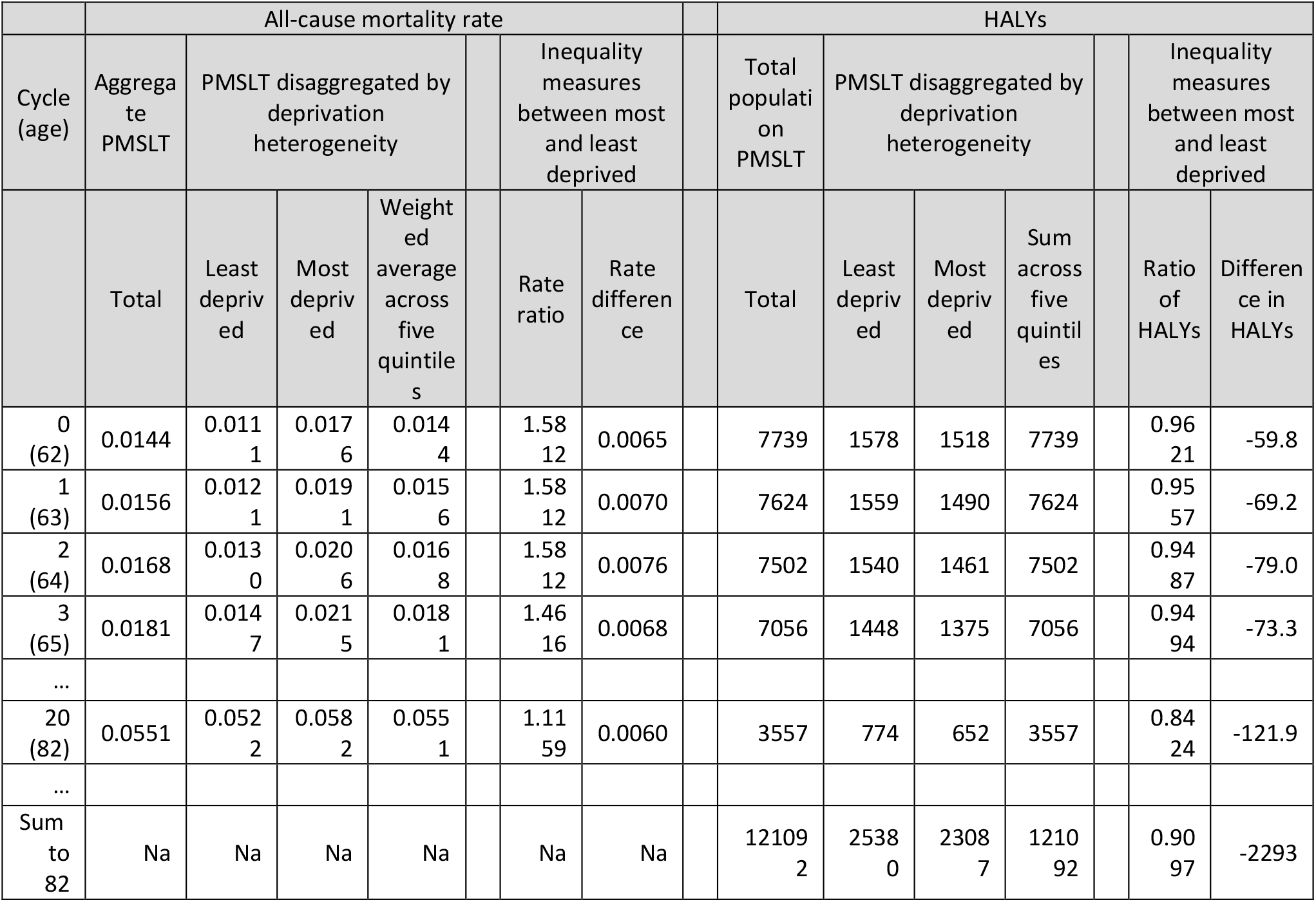
Business as usual 60-64 year-old cohort (centered on age 62) of MĀori females, comparing aggregated and deprivation heterogeneity disaggregated proportional multistate lifetables (PMSLT) models in term of all-cause mortality rates and health adjusted life years (HALYs) outputs †

Table 2 shows the three intervention scenarios. For the (extreme) scenario of 50% reductions in ACMRs at all ages, the rate ratios comparing the most and least deprived Māori females are unchanged (as per specification), and the rate differences halved. By the age of 82, there is a difference in the aggregated population mortality rate (0.0275) and that averaged (weighted) across quintiles (0.0276) at the third meaningful digit. Similarly, summed to age 82 the total HALYs <u>incremental to BAU</u> differ by 30 (0.2%) for the aggregate (13233) compared to heterogeneity (13204) models – due to the non-linear association of mortality rates with mortality risks, with mortality rates varying by strata.

**Table 2:** Intervention impacts for 60-64 year-old cohort of MĀori females, comparing aggregated and deprivation heterogeneity disaggregated proportional multistate lifetables (PMSLT) models in terms of all-cause mortality rates and health adjusted life years (HALYs) outputs

|  | All-cause mortality rate |  |  |  |  |  |  |  | HALYs <u>incremental to BAU</u> |  |  |  |  |  |  |
| --- | --- | --- | --- | --- | --- | --- | --- | --- | --- | --- | --- | --- | --- | --- | --- |
| Cycle (age) | Aggreg<br>ate<br>PMSLT | PMSLT disaggregated<br>by deprivation<br>heterogeneity |  |  |  | Inequality measures<br>between most and<br>least deprived (% shift<br>to null c.f. BAU [Table<br>1]) |  |  | Aggreg<br>ate<br>PMSLT | PMSLT disaggregated<br>by deprivation<br>heterogeneity |  |  |  | Inequality<br>most: least<br>deprived |  |
|  | Total | Least<br>depriv<br>ed | Most<br>depriv<br>ed | Avera<br>ge<br>across<br>five<br>quintil<br>es |  | Rate ratio | Rate<br>difference |  | Total | Least<br>depriv<br>ed | Most<br>depriv<br>ed | Sum<br>across<br>quintil<br>es |  | Rati<br>o of<br>HALY<br>s | Differ<br>ence in<br>HALYs |
| Intervention A: 50% reduction in ACMR rate |  |  |  |  |  |  |  |  |  |  |  |  |  |  |  |
| 0<br>(62) | 0.0072 | 0.0056 | 0.0088 | 0.0072 |  | 1.581<br>(0%) | 0.0032<br>(50%) |  | 28 | 4 | 7 | 28 |  | 1.52 | 2.3 |
| 1<br>(63) | 0.0078 | 0.0060 | 0.0095 | 0.0078 |  | 1.581<br>(0%) | 0.0035<br>(50%) |  | 85 | 13 | 20 | 85 |  | 1.51 | 6.9 |
| 2<br>(64) | 0.0084 | 0.0065 | 0.0103 | 0.0084 |  | 1.581<br>(0%) | 0.0038<br>(50%) |  | 145 | 23 | 35 | 145 |  | 1.51 | 11.7 |
| 3<br>(65) | 0.0091 | 0.0074 | 0.0108 | 0.0091 |  | 1.462<br>(0%) | 0.0034<br>(50%) |  | 200 | 32 | 48 | 199 |  | 1.49 | 15.7 |
| 20<br>(82) | 0.0275 | 0.0261 | 0.0291 | 0.0276 |  | 1.116<br>(0%) | 0.0030<br>(50%) |  | 1178 | 220 | 249 | 1175 |  | 1.13 | 29.0 |
| Sum<br>to<br>82 |  |  |  |  |  |  |  |  | 13233 | 2325 | 2939 | 13204 |  | 1.26 | 614 |
| Intervention B: 50% reduction in CHD incidence rate |  |  |  |  |  |  |  |  |  |  |  |  |  |  |  |
| 0<br>(62) | 0.0144 | 0.0111 | 0.0176 | 0.0144 |  | 1.581<br>(0.0%) | 0.0065<br>(0.0%) |  | 3.0 | 0.5 | 0.7 | 3.0 |  | 1.62 | 0.28 |
| 1<br>(63) | 0.0155 | 0.0120 | 0.0190 | 0.0155 |  | 1.579<br>(0.4%) | 0.0070<br>(0.9%) |  | 9.3 | 1.4 | 2.3 | 9.3 |  | 1.62 | 0.87 |
| 2<br>(64) | 0.0166 | 0.0129 | 0.0204 | 0.0166 |  | 1.577<br>(0.7%) | 0.0075<br>(1.7%) |  | 16.5 | 2.5 | 4.0 | 16.4 |  | 1.61 | 1.52 |
| 3<br>(65) | 0.0178 | 0.0145 | 0.0211 | 0.0177 |  | 1.455<br>(1.5%) | 0.0066<br>(2.9%) |  | 21.7 | 3.3 | 5.2 | 21.7 |  | 1.59 | 1.92 |
| 20<br>(82) | 0.0504 | 0.0482 | 0.0529 | 0.0504 |  | 1.099<br>(14.9%) | 0.0048<br>(21.4%) |  | 169 | 30 | 36 | 168 |  | 1.20 | 6.11 |
| Sum<br>to<br>82 |  |  |  |  |  |  |  |  | 1697 | 283 | 378 | 1688 |  | 1.34 | 94.8 |
| Intervention C: 59% sodium chloride substitution with potassium chloride |  |  |  |  |  |  |  |  |  |  |  |  |  |  |  |
| 0<br>(62) | 0.0144 | 0.0111 | 0.0176 | 0.0144 |  | 1.581<br>(0.0%) | 0.0065<br>(0.0%) |  | 0.51 | 0.08 | 0.12 | 0.50 |  | 1.63 | 0.05 |
| 1<br>(63) | 0.0156 | 0.0121 | 0.0191 | 0.0156 |  | 1.581<br>(0.1%) | 0.0070<br>(0.2%) |  | 1.58 | 0.24 | 0.39 | 1.57 |  | 1.63 | 0.15 |
| 2<br>(64) | 0.0168 | 0.0130 | 0.0206 | 0.0168 |  | 1.580<br>(0.1%) | 0.0076<br>(0.3%) |  | 2.77 | 0.41 | 0.67 | 2.77 |  | 1.62 | 0.26 |
| 3<br>(65) | 0.0180 | 0.0147 | 0.0215 | 0.0180 |  | 1.460<br>(0.3%) | 0.0068<br>(0.5%) |  | 3.85 | 0.58 | 0.92 | 3.84 |  | 1.60 | 0.35 |
| Cycle (age) | Aggregate PMSLT | PMSLT disaggregated by deprivation heterogeneity |  |  |  | Inequality measures between most and least deprived (% shift to null c.f. BAU [Table 1]) |  |  | Aggregate PMSLT | PMSLT disaggregated by deprivation heterogeneity |  |  |  | Inequality most: least deprived |  |
|  | Total | Least deprived | Most deprived | Average across five quintiles |  | Rate ratio | Rate difference |  | Total | Least deprived | Most deprived | Sum across quintiles |  | Ratio of HALYs | Difference in HALYs |
| 20 (82) | 0.0545 | 0.0517 | 0.0576 | 0.0545 |  | 1.114 (1.5%) | 0.0059 (2.4%) |  | 21.9 | 4.0 | 4.6 | 21.7 |  | 1.17 | 0.67 |
| Sum to 82 |  |  |  |  |  |  |  |  | 229.9 | 38.5 | 51.0 | 228.7 |  | 1.32 | 12.51 |

Also shown in Table 2 are the measures of interest to assessing health inequality impacts of the interventions. For an intervention reducing CHD incidence by 50% and a ‘real world’ scenario of sodium substitution reducing both CHR and stroke incidence, there are reductions to the null in the ACMR rate ratio and rate differences – due to the higher rates of these diseases in deprived populations that makes these interventions inequality reducing. Similarly, there are greater HALY gains for the deprived population in both relative and absolute terms (last two columns of Table 2).

## Discussion

We developed algorithms for mathematically disaggregating a population into strata such that the lifetables for the sub-populations under BAU are consistent with the lifetable of the aggregate population. To the best of our knowledge, this method is the first of its kind to perform this disaggregation with mathematical guarantees of consistency for the sub-populations (see discussion in Section 1). These guarantees allow the lifetables for the sub-populations to be treated as the “working truth” and thus they can be used in simulations to estimate HALY gains by strata without any loss of fidelity.

We find a modest difference in the sum of health adjusted life years (HALYs) gained across strata compared to ‘simply’ applying the intervention to the aggregate population – a difference that arises due to the non-linear association of rates with risks (i.e. risk = 1 – exp[rate]), and where the intervention effect modeled separately across strata then summed is more accurate than modeled simply in aggregate (as heterogeneity is allowed for).

There are two main assumptions that are required for the disaggregated outputs to be reliable. First, we assume that the lifetable for the aggregate population has been correctly parameterized and that the outputs of the BAU simulations for the aggregate lifetables are accurate to for the population they model. This may be a strong assumption in practice; lifetable parameters are often obtained through projections and approximations and may involve a significant degree of uncertainty. However, since the sub-populations are mathematically consistent with the aggregate population, the amount of error in the estimated sub-population is only as large as that of the aggregate.

The second assumption is that the rate ratios comparing strata (e.g. disease incidence rate ratios), and the initial proportions in each sub-population accurately reflect reality. This implies that one must have accurate *a priori* knowledge of the relative differences between sub-populations.

The disaggregation algorithm is not computationally expensive to implement for typical applications (i.e., a manageable number of sub-populations), and should assist researchers aiming to quantify intervention effects by population heterogeneity.

## Data Availability

All of the data used in this study were sourced from previously published research by the authors.

## Appendix 1

### Abstracts from Literature Search

We conducted a literature review to determine if methods to solve the problem of consistently solving for sub-populations with simulation models had previously been proposed. We used the PubMed search engine with the Boolean string “(population) AND (disaggregation OR heterogeneity) AND (life table OR Markov process OR Markov chain) AND (rate OR transition probability) AND (technique OR algorithm)”, which was automatically expanded on by the engine to include related and MeSH terms. This returned 82 results, to which we added 5 articles that were found through previous searches. After screening titles and abstracts for relevancy, the list of articles was reduced to 14 (see below), of which we determined that none involved the same problem as investigated in this paper.

1. **Title: The burden of chronic respiratory diseases and their heterogeneity across the states of India: the Global Burden of Disease Study 1990-2016**. Abstract: BACKGROUND: India has 18% of the global population and an increasing burden of chronic respiratory diseases. However, a systematic understanding of the distribution of chronic respiratory diseases and their trends over time is not readily available for all of the states of India. Our aim was to report the trends in the burden of chronic respiratory diseases and the heterogeneity in their distribution in all states of India between 1990 and 2016. METHODS: Using all accessible data from multiple sources, we estimated the prevalence of major chronic respiratory diseases and the deaths and disability-adjusted life-years (DALYs) caused by them for every state of India from 1990 to 2016 as part of the Global Burden of Diseases, Injuries, and Risk Factors Study (GBD) 2016. We assessed heterogeneity in the burden of chronic obstructive pulmonary disease (COPD) and asthma across the states of India. The states were categorised into four groups based on their epidemiological transition level (ETL). ETL was defined as the ratio of DALYs from communicable diseases to those from non-communicable diseases and injuries combined, with a low ratio denoting high ETL and vice versa. We also assessed the contribution of risk factors to DALYs due to COPD. We compared the burden of chronic respiratory diseases in India against the global average in GBD 2016. We calculated 95% uncertainty intervals (UIs) for the point estimates. FINDINGS: The contribution of chronic respiratory diseases to the total DALYs in India increased from 4·5% (95% UI 4·0-4·9) in 1990 to 6·4% (5·8-7·0) in 2016. Of the total global DALYs due to chronic respiratory diseases in 2016, 32·0% occurred in India. COPD and asthma were responsible for 75·6% and 20·0% of the chronic respiratory disease DALYs, respectively, in India in 2016. The number of cases of COPD in India increased from 28·1 million (27·0-29·2) in 1990 to 55·3 million (53·1-57·6) in 2016, an increase in prevalence from 3·3% (3·1-3·4) to 4·2% (4·0-4·4). The age-standardised COPD prevalence and DALY rates in 2016 were highest in the less developed low ETL state group. There were 37·9 million (35·7-40·2) cases of asthma in India in 2016, with similar prevalence in the four ETL state groups, but the highest DALY rate was in the low ETL state group. The highest DALY rates for both COPD and asthma in 2016 were in the low ETL states of Rajasthan and Uttar Pradesh. The DALYs per case of COPD and asthma were 1·7 and 2·4 times higher in India than the global average in 2016, respectively; most states had higher rates compared with other locations worldwide at similar levels of Socio-demographic Index. Of the DALYs due to COPD in India in 2016, 53·7% (43·1-65·0) were attributable to air pollution, 25·4% (19·5-31·7) to tobacco use, and 16·5% (14·1-19·2) to occupational risks, making these the leading risk factors for COPD. INTERPRETATION: India has a disproportionately high burden of chronic respiratory diseases. The increasing contribution of these diseases to the overall disease burden across India and the high rate of health loss from them, especially in the less developed low ETL states, highlights the need for focused policy interventions to address this significant cause of disease burden in India. FUNDING: Bill &Melinda Gates Foundation; and Indian Council of Medical Research, Department of Health Research, Ministry of Health and Family Welfare, Government of India Reference: (18)
2. **Title: Interacting effects of unobserved heterogeneity and individual stochasticity in the life history of the southern fulmar**. Abstract: Individuals are heterogeneous in many ways. Some of these differences are incorporated as individual states (e.g. age, size, breeding status) in population models. However, substantial amounts of heterogeneity may remain unaccounted for, due to unmeasurable genetic, maternal or environmental factors. Such unobserved heterogeneity (UH) affects the behaviour of heterogeneous cohorts via intra-cohort selection and contributes to inter-individual variance in demographic outcomes such as longevity and lifetime reproduction. Variance is also produced by individual stochasticity, due to random events in the life cycle of wild organisms, yet no study thus far has attempted to decompose the variance in demographic outcomes into contributions from UH and individual stochasticity for an animal population in the wild. We developed a stage-classified matrix population model for the southern fulmar breeding on Ile des Pétrels, Antarctica. We applied multievent, multistate mark-recapture methods to estimate a finite mixture model accounting for UH in all vital rates and Markov chain methods to calculate demographic outcomes. Finally, we partitioned the variance in demographic outcomes into contributions from UH and individual stochasticity. We identify three UH groups, differing substantially in longevity, lifetime reproductive output, age at first reproduction and in the proportion of the life spent in each reproductive state. -14% of individuals at fledging have a delayed but high probability of recruitment and extended reproductive life span. -67% of individuals are less likely to reach adulthood, recruit late and skip breeding often but have the highest adult survival rate. -19% of individuals recruit early and attempt to breed often. They are likely to raise their offspring successfully, but experience a relatively short life span. Unobserved heterogeneity only explains a small fraction of the variances in longevity (5.9%), age at first reproduction (3.7%) and lifetime reproduction (22%). UH can affect the entire life cycle, including survival, development and reproductive rates, with consequences over the lifetime of individuals and impacts on cohort dynamics. The respective role of UH vs. individual stochasticity varies greatly among demographic outcomes. We discuss the implication of our finding for the gradient of life-history strategies observed among species and argue that individual differences should be accounted for in demographic studies of wild populations. Reference: (19)
3. **Title: The impact of individual-level heterogeneity on estimated infectious disease burden: a simulation study**. Abstract: BACKGROUND: Disease burden is not evenly distributed within a population; this uneven distribution can be due to individual heterogeneity in progression rates between disease stages. Composite measures of disease burden that are based on disease progression models, such as the disability-adjusted life year (DALY), are widely used to quantify the current and future burden of infectious diseases. Our goal was to investigate to what extent ignoring the presence of heterogeneity could bias DALY computation. METHODS: Simulations using individual-based models for hypothetical infectious diseases with short and long natural histories were run assuming either “population-averaged” progression probabilities between disease stages, or progression probabilities that were influenced by an a priori defined individual-level frailty (i.e., heterogeneity in disease risk) distribution, and DALYs were calculated. RESULTS: Under the assumption of heterogeneity in transition rates and increasing frailty with age, the short natural history disease model predicted 14% fewer DALYs compared with the homogenous population assumption. Simulations of a long natural history disease indicated that assuming homogeneity in transition rates when heterogeneity was present could overestimate total DALYs, in the present case by 4% (95% quantile interval: 1-8%). CONCLUSIONS: The consequences of ignoring population heterogeneity should be considered when defining transition parameters for natural history models and when interpreting the resulting disease burden estimates. Reference: (20)
4. **Title: Estimation of heterogeneity in malaria transmission by stochastic modelling of apparent deviations from mass action kinetics**. Abstract: BACKGROUND: Quantifying heterogeneity in malaria transmission is a prerequisite for accurate predictive mathematical models, but the variance in field measurements of exposure overestimates true micro-heterogeneity because it is inflated to an uncertain extent by sampling variation. Descriptions of field data also suggest that the rate of Plasmodium falciparum infection is not proportional to the intensity of challenge by infectious vectors. This appears to violate the principle of mass action that is implied by malaria biology. Micro-heterogeneity may be the reason for this anomaly. It is proposed that the level of micro-heterogeneity can be estimated from statistical models that estimate the amount of variation in transmission most compatible with a mass-action model for the relationship of infection to exposure. METHODS: The relationship between the entomological inoculation rate (EIR) for falciparum malaria and infection risk was reanalysed using published data for cohorts of children in Saradidi (western Kenya). Infection risk was treated as binomially distributed, and measurement-error (Poisson and negative binomial) models were considered for the EIR. Models were fitted using Bayesian Markov chain Monte Carlo algorithms and model fit compared for models that assume either mass-action kinetics, facilitation, competition or saturation of the infection process with increasing EIR. RESULTS: The proportion of inocula that resulted in infection in Saradidi was inversely related to the measured intensity of challenge. Models of facilitation showed, therefore, a poor fit to the data. When sampling error in the EIR was neglected, either competition or saturation needed to be incorporated in the model in order to give a good fit. Negative binomial models for the error in exposure could achieve a comparable fit while incorporating the more parsimonious and biologically plausible mass action assumption. Models that assume negative binomial micro-heterogeneity predict lower incidence of infection at a given average exposure than do those assuming exposure to be uniform. The negative binomial model moreover provides an estimate of the variance of the within-cohort distribution of the EIR and hence of within cohort heterogeneity in exposure. CONCLUSION: Apparent deviations from mass action kinetics in parasite transmission can arise from spatial and temporal heterogeneity in the inoculation rate, and from imprecision in its measurement. For parasites like P. falciparum, where there is no plausible biological rationale for deviations from mass action, this provides a strategy for estimating true levels of heterogeneity, since if mass-action is assumed, the within-population variance in exposure becomes identifiable in cohort studies relating infection to transmission intensity. Statistical analyses relating infection to exposure thus provide a valid general approach for estimating heterogeneity in transmission but only when they incorporate mass action kinetics and shrinkage estimates of exposure. Such analyses make it possible to include realistic levels of heterogeneity in dynamic models that predict the impact of control measures on transmission intensity. Reference: (21)
5. **Title: A class of latent Markov models for capture-recapture data allowing for time, heterogeneity, and behavior effects**. Abstract: We propose an extension of the latent class model for the analysis of capture-recapture data which allows us to take into account the effect of a capture on the behavior of a subject with respect to future captures. The approach is based on the assumption that the variable indexing the latent class of a subject follows a Markov chain with transition probabilities depending on the previous capture history. Several constraints are allowed on these transition probabilities and on the parameters of the conditional distribution of the capture configuration given the latent process. We also allow for the presence of discrete explanatory variables, which may affect the parameters of the latent process. To estimate the resulting models, we rely on the conditional maximum likelihood approach and for this aim we outline an EM algorithm. We also give some simple rules for point and interval estimation of the population size. The approach is illustrated by applying it to two data sets concerning small mammal populations. Reference: (22)
6. **Title: Analysis of the relationship between socioeconomic factors and stomach cancer incidence in Slovenia**. Abstract: An unequal population distribution of well-known major risk factors explains much of the variation in the incidence of stomach cancer worldwide. The aim of this study was to determine whether geographical variation of the stomach cancer incidence rate between Slovenia’s municipalities during years 1995-2001 could partially be explained by variations in the socioeconomic status as an indirect stomach cancer risk factor. A composite measure of each region’s socioeconomic status, labelled as deprivation index, was created from basic socioeconomic characteristics of each municipality using factor analysis. Municipalities’ standardized incidence ratios for all stomach cancers and non-cardia stomach cancer were calculated. A fully Bayesian spatial model with a conditionally autoregressive prior was applied using Markov chain Monte Carlo techniques and WinBUGS software. Spatially smoothed maps of stomach cancer incidence rates by 192 Slovenian municipalities show a clear west-to-east gradient. This pattern resembles the geographical variation of socioeconomic indices, but these indices are not significant predictors of stomach cancer incidence. Geographical variation of stomach cancer incidence in Slovenia could be partially explained by the heterogeneous socioeconomic characteristics of its municipalities. It is possible that the socioeconomic status indices used in our study were not enough powerful predictors of stomach cancer risk. Some further methodological research is needed to explain why this association was not statistically evident with the current modeling approach. Reference: (23)
7. **Title: Calculation of disease dynamics in a population of households**. Abstract: Early mathematical representations of infectious disease dynamics assumed a single, large, homogeneously mixing population. Over the past decade there has been growing interest in models consisting of multiple smaller subpopulations (households, workplaces, schools, communities), with the natural assumption of strong homogeneous mixing within each subpopulation, and weaker transmission between subpopulations. Here we consider a model of SIRS (susceptible-infectious-recovered-susceptible) infection dynamics in a very large (assumed infinite) population of households, with the simplifying assumption that each household is of the same size (although all methods may be extended to a population with a heterogeneous distribution of household sizes). For this households model we present efficient methods for studying several quantities of epidemiological interest: (i) the threshold for invasion; (ii) the early growth rate; (iii) the household offspring distribution; (iv) the endemic prevalence of infection; and (v) the transient dynamics of the process. We utilize these methods to explore a wide region of parameter space appropriate for human infectious diseases. We then extend these results to consider the effects of more realistic gamma-distributed infectious periods. We discuss how all these results differ from standard homogeneous-mixing models and assess the implications for the invasion, transmission and persistence of infection. The computational efficiency of the methodology presented here will hopefully aid in the parameterisation of structured models and in the evaluation of appropriate responses for future disease outbreaks. Reference: (24)
8. **Title: Estimating heterogeneous transmission with multiple infectives using MCMC methods**. Abstract: We developed a general procedure for estimating the transmission probability adjusting for covariates when susceptibles are exposed to several infectives concurrently and taking correlation within transmission units into account. The procedure is motivated by a study estimating efficacy of pertussis vaccination based on the secondary attack rate in a rural sub-Saharan community (Niakhar, Senegal) and illustrated with simulations. The procedure is also appropriate to estimate the pairwise transmission probability in transmission studies of live vaccine virus in a collection of transmission units, such as day-care centres or retirement centres. Previously, analyses either excluded transmission units with multiple infectives or ignored co-infectives. Excluding transmission units with multiple infectives is statistically less efficient and ignoring co-infectives can lead to biased estimation. Modelling is carried out by regressing the latent pairwise transmission probability from each infective to a susceptible on covariates and specifying a transmission linkage function linking the latent pairwise transmission probability to the overall transmission probability. Parameters are estimated using Markov chain Monte Carlo methods. Reference: (25)
9. **Title: Mortality of a heterogeneous cohort; description and implications**. Abstract: A recent model for heterogeneous mortality by Vaupel et al. is shown to be based on incorrect definitions. An alternative formulation is presented. The results indicate that current methods for computing the survivorship and life expectation functions underestimate the true values. A method is given for determining the possible magnitude of this underestimation. The method is illustrated by a numerical example using U.S. data. Reference: (26)
10. **Title: A New Approach to Estimating Life Tables with Covariates and Constructing Interval Estimates of Life Table Quantities**. Abstract: Extant approaches to constructing life tables generally rely on the use of population data, and differences between groups defined by discrete characteristics are examined by disaggregating the data before estimation. When sample data are used, few researchers have attempted to include covariates directly in the process of estimation, and fewer still have attempted to construct interval estimates for state expectancies when covariates are used. In this paper, we present a Bayesian approach that is useful for producing interval estimates for single-decrement, multiple-decrement, and multistate life tables. The method involves (1) estimating a hazard or survival model using Bayesian Markov chain Monte Carlo (MCMC) methods to produce a sample from the posterior distribution for the parameters of the model; (2) generating distributions of transition probabilities for selected values of covariates using the sample of model parameters; (3) using these distributions of transition probabilities as inputs for life table construction; and (4) summarizing the distribution of life table quantities. We illustrate the method on data simulated from the Berkeley Mortality Database, data from the National Health and Nutrition Examination Survey (and follow-ups), and data from the National Long Term Care Survey, and we show how the results can be used for hypothesis testing. Reference: (27)
11. **Title: Disaggregation of Statistical Livestock Data Using the Entropy Approach**. Abstract: A process of agricultural data disaggregation is developed to address the lack of updated disaggregated data concerning main livestock categories at subregional and county level in the Alentejo Region, southern Portugal. The model developed considers that the number of livestock units is a function of the agricultural and forest occupation, and data concerning the existing agricultural and forest occupation, as well as the conversion of livestock numbers into normal heads, are needed in order to find this relation. The weight of each livestock class is estimated using a dynamic process based on a generalized maximum entropy model and on a crossentropy minimization model, which comprises two stages. The model was applied to the county of Castelo de Vide and their results were validated in cross reference to real data from different sources. Reference: (28)
12. **Title: Life table methods for heterogeneous populations: distributions describing the heterogeneity**. Abstract: Taking account of heterogeneity between the individuals in population based mortality studies is important. A systematic way of describing heterogeneity is by an unobserved quantity called frailty, entering the hazard multiplicatively. Until now most studies have used a gamma distributed frailty, which is mathematically convenient; for example, the distribution among survivors is also gamma. This paper shows that several other distributions have equally simple properties, the main example being the inverse Gaussian distribution. Consequences of the different distributions are examined; the inverse Gaussian makes the population homogeneous with time, whereas for the gamma the relative heterogeneity is constant. Reference: (29)
13. **Title: Obtaining Multistate Life Table Distributions for Highly Refined Subpopulations From Cross-Sectional Data: A Bayesian Extension of Sullivan’s Method**. Abstract: Multistate life table methods are often used to estimate the proportion of remaining life that individuals can expect to spend in various states, such as healthy and unhealthy states. Sullivan’s method is commonly used when panels containing data on transitions are unavailable and true multistate tables cannot be generated. Sullivan’s method requires only cross-sectional mortality data and cross-sectional data indicating prevalence in states of interest. Such data often come from sample surveys, which are widely available. Although the data requirements for Sullivan’s method are minimal, the method is limited in its ability to produce estimates for subpopulations because of limited disaggregation of data in cross-sectional mortality files and small cell sizes in aggregated survey data. In this article, we develop, test, and demonstrate a method that adapts Sullivan’s approach to allow the inclusion of covariates in producing interval estimates of state expectancies for any desired subpopulation that can be specified in the cross-sectional prevalence data. The method involves a three-step process: (1) using Gibbs sampling to sample parameters from a bivariate regression model; (2) using ecological inference for producing transition probability matrices from the Gibbs samples; (3) using standard multistate calculations to convert the transition probability matrices into multistate life tables. Reference: (10)
14. **Title: On the heterogeneity of human populations as reflected by mortality dynamics**. Abstract: The heterogeneity of populations is used to explain the variability of mortality rates across the lifespan and their deviations from an exponential growth at young and very old ages. A mathematical model that combines the heterogeneity with the assumption that the mortality of each constituent subpopulation increases exponentially with age, has been shown to successfully reproduce the entire mortality pattern across the lifespan and its evolution over time. In this work we aim to show that the heterogeneity is not only a convenient consideration for fitting mortality data but is indeed the actual structure of the population as reflected by the mortality dynamics over age and time. In particular, we show that the model of heterogeneous population fits mortality data better than other commonly used mortality models. This was demonstrated using cohort data taken for the entire lifespan as well as for only old ages. Also, we show that the model can reproduce seemingly contradicting observations in late-life mortality dynamics. Finally, we show that the homogenisation of a population, observed by fitting the model to actual data of consecutive periods, can be associated with the evolution of allele frequencies if the heterogeneity is assumed to reflect the genetic variations within the population. Reference: (30)

## Appendix 2

### Disaggregation Details

Given mortality rate ratios 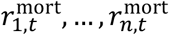 for timestep *t* with 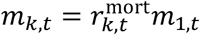 for each sub-population *k*, our goal is to find sub-population mortality rates *m*_*k,t*_ such that:

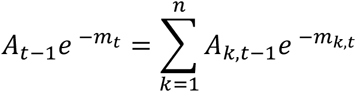

for each timestep *t*. By substituting 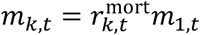 into the constraint, we obtain the equation:

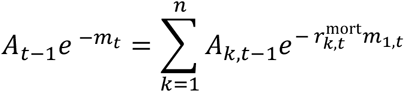

Consider the constraint for the first timestep, where *t* = 1.

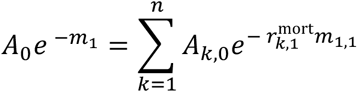

Let 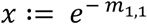. If we assume that *m*_1,1_ ≥ 0, then 0 < *x* ≤ 1. By substituting *x* into the equation, we obtain:

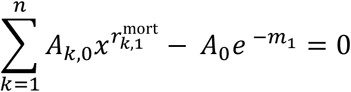

Let 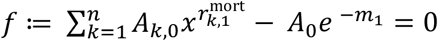. Assuming that *A* ≥ 0 for each *k*, and 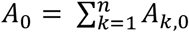, it can be seen that *ff*(0) < 0 and *f*(1) ≥ 1. Since *f* is monotonically increasing in the interval (0, 1], this implies that *f* has a unique root for *x* ∈ (0, 1]. Let *x*′ be the unique root for *f* in the interval (0, 1]. Then *m*′_1,1_ := − log(*x*′) is the unique rate in the interval [0, ∞) that satisfies the constraint. By using the rate ratios, we can obtain mortality rates for each sub-population at time *t* = 1 in terms of *m*′_1,1_ such that the sub-population quantities for the alive compartment are consistent with the aggregate population at *t* = 1. In order to obtain a value for *x*′, one can use one of several root finding methods to obtain a root for *f*(*x*) in the interval (0, 1], such as Brent’s method (31).

Once the sub-population mortality rates are known for *t* = 1, we can use the recursive formulae for the alive and dead populations to obtain explicit values for the population quantities *A*_*k*,1_ and *D*_*k*,1_. The above process can then be repeated for *t* = 2 with the constraint

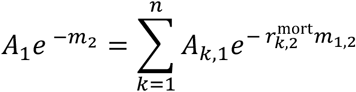

to obtain the unique rate *m*′_1,2_ which satisfies the constraint. By continuing to iterate this process, we can compute the sub-population mortality rates and population quantities up to any time period.

We summarize this procedure as the following algorithm.

Mortality Lifetable Disaggregation Algorithm

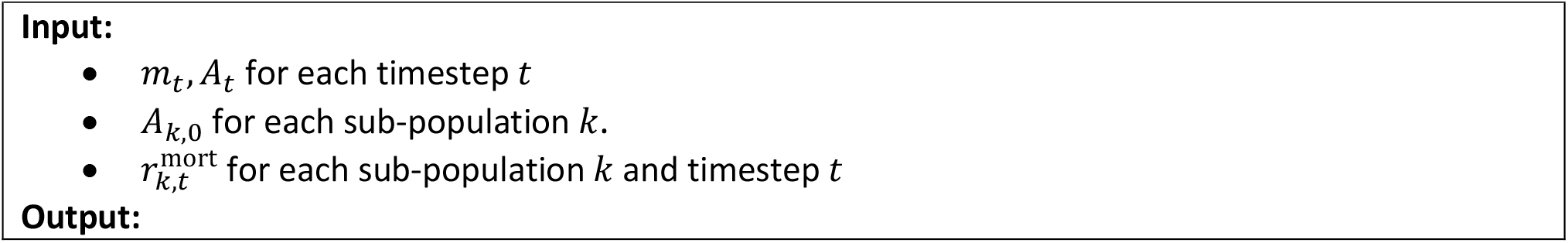
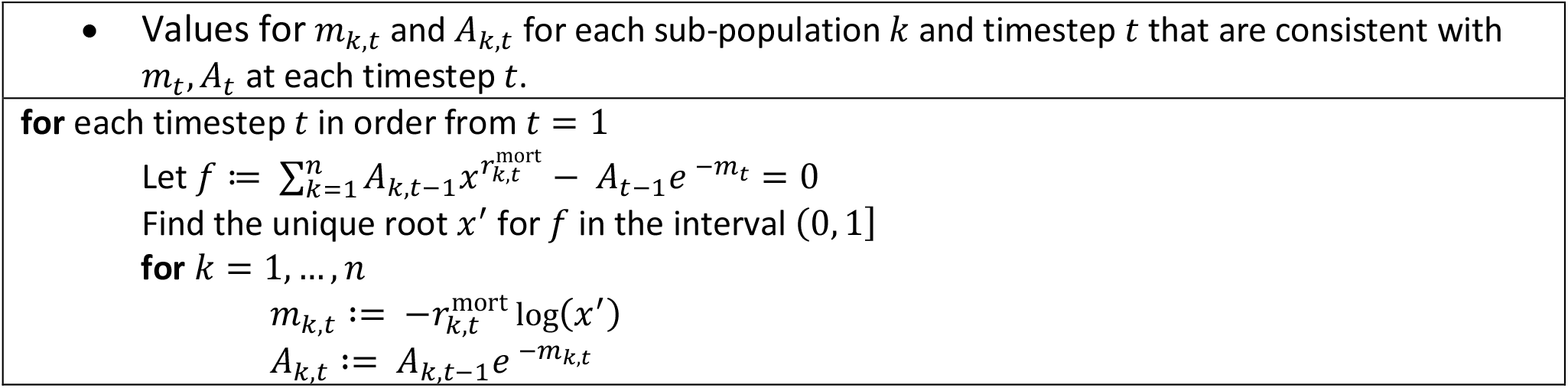

## Appendix 2

**Supplementary Table 1:**
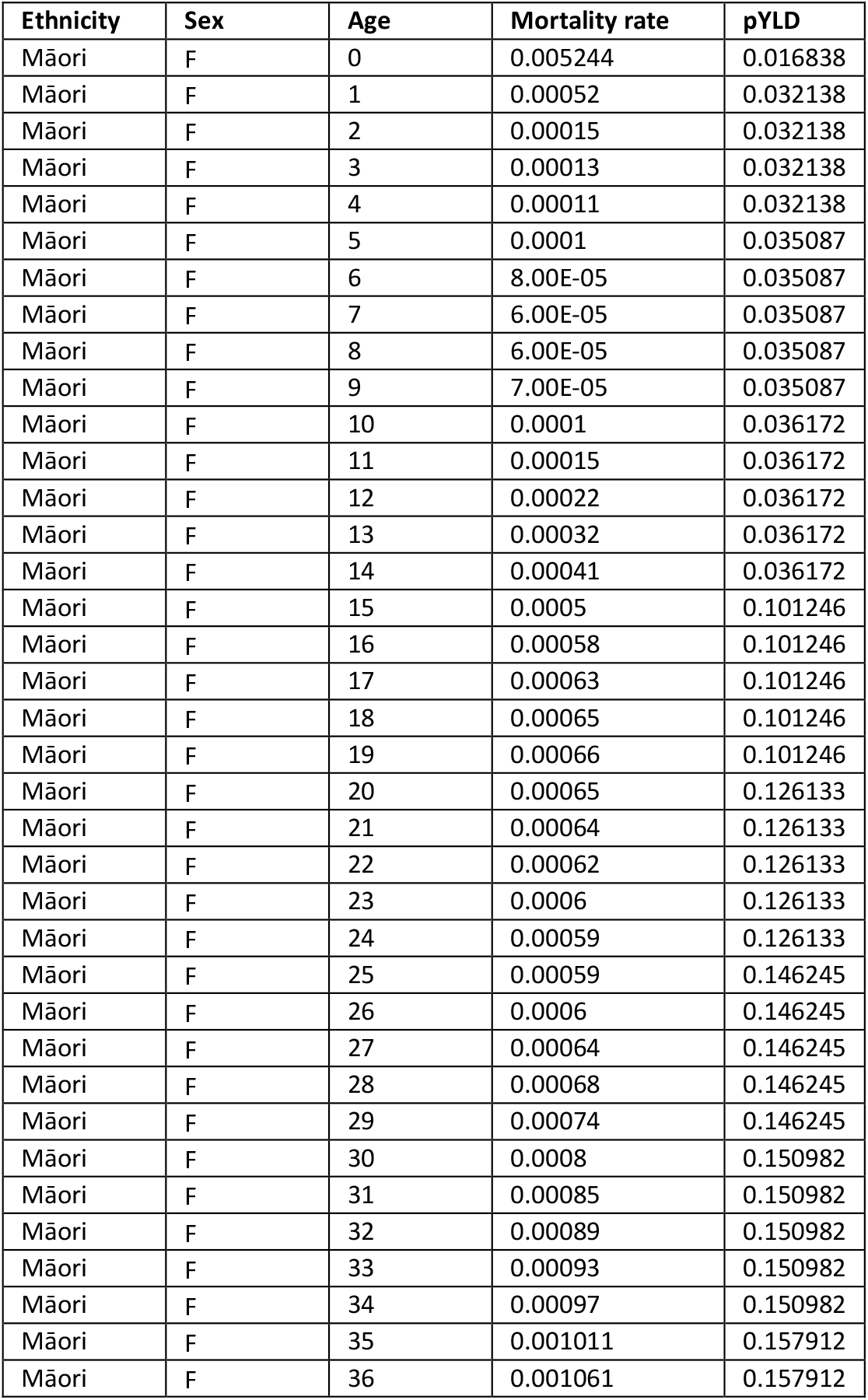
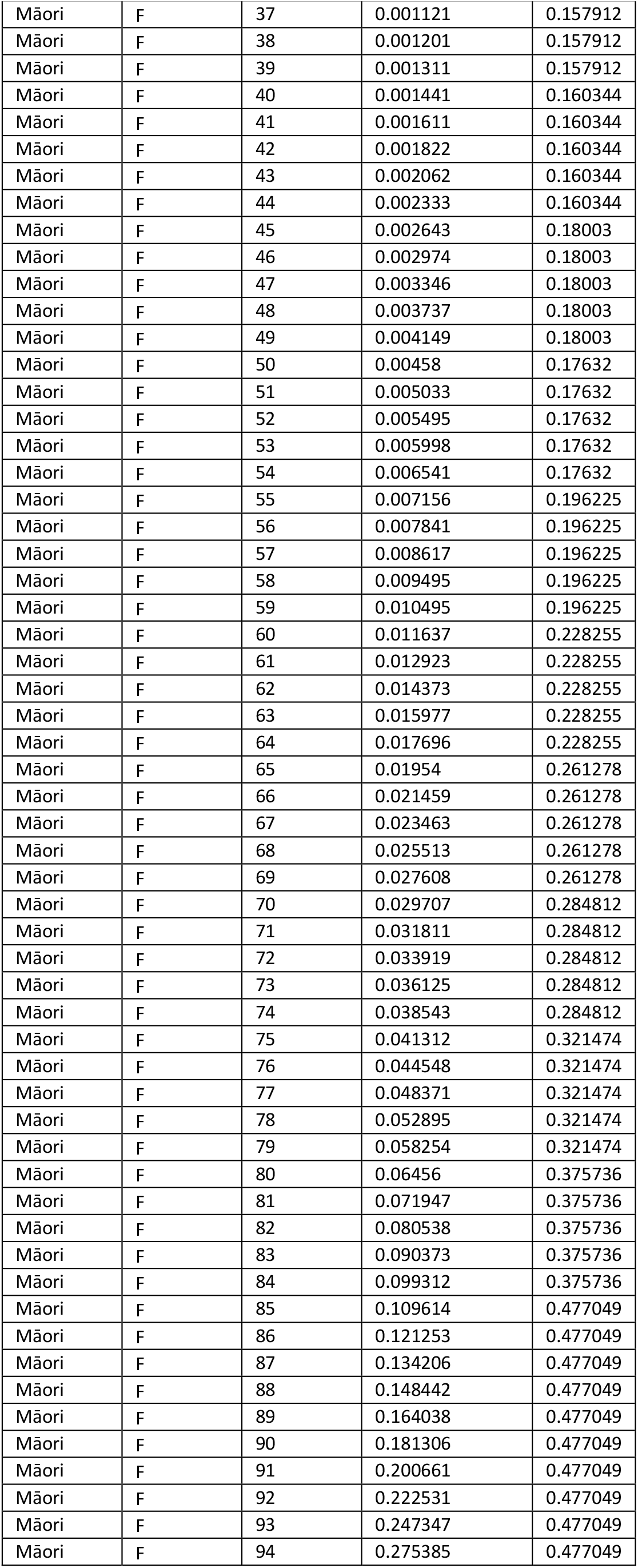
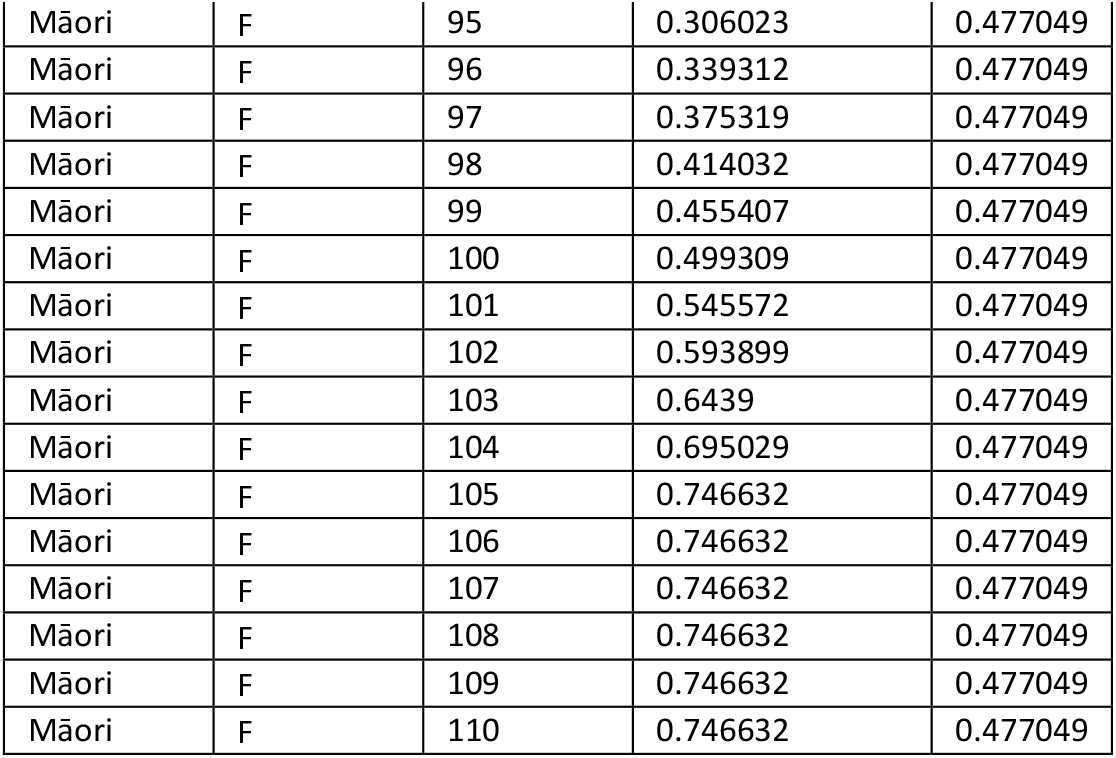
Base Mortality rate and pYLD for MĀori Females

| Ethnicity | Sex | Age | Mortality rate | pYLD |
| --- | --- | --- | --- | --- |
| Māori | F | 0 | 0.005244 | 0.016838 |
| Māori | F | 1 | 0.00052 | 0.032138 |
| Māori | F | 2 | 0.00015 | 0.032138 |
| Māori | F | 3 | 0.00013 | 0.032138 |
| Māori | F | 4 | 0.00011 | 0.032138 |
| Māori | F | 5 | 0.0001 | 0.035087 |
| Māori | F | 6 | 8.00E-05 | 0.035087 |
| Māori | F | 7 | 6.00E-05 | 0.035087 |
| Māori | F | 8 | 6.00E-05 | 0.035087 |
| Māori | F | 9 | 7.00E-05 | 0.035087 |
| Māori | F | 10 | 0.0001 | 0.036172 |
| Māori | F | 11 | 0.00015 | 0.036172 |
| Māori | F | 12 | 0.00022 | 0.036172 |
| Māori | F | 13 | 0.00032 | 0.036172 |
| Māori | F | 14 | 0.00041 | 0.036172 |
| Māori | F | 15 | 0.0005 | 0.101246 |
| Māori | F | 16 | 0.00058 | 0.101246 |
| Māori | F | 17 | 0.00063 | 0.101246 |
| Māori | F | 18 | 0.00065 | 0.101246 |
| Māori | F | 19 | 0.00066 | 0.101246 |
| Māori | F | 20 | 0.00065 | 0.126133 |
| Māori | F | 21 | 0.00064 | 0.126133 |
| Māori | F | 22 | 0.00062 | 0.126133 |
| Māori | F | 23 | 0.0006 | 0.126133 |
| Māori | F | 24 | 0.00059 | 0.126133 |
| Māori | F | 25 | 0.00059 | 0.146245 |
| Māori | F | 26 | 0.0006 | 0.146245 |
| Māori | F | 27 | 0.00064 | 0.146245 |
| Māori | F | 28 | 0.00068 | 0.146245 |
| Māori | F | 29 | 0.00074 | 0.146245 |
| Māori | F | 30 | 0.0008 | 0.150982 |
| Māori | F | 31 | 0.00085 | 0.150982 |
| Māori | F | 32 | 0.00089 | 0.150982 |
| Māori | F | 33 | 0.00093 | 0.150982 |
| Māori | F | 34 | 0.00097 | 0.150982 |
| Māori | F | 35 | 0.001011 | 0.157912 |
| Māori | F | 36 | 0.001061 | 0.157912 |

| **Ethnicity** | **Sex** | **Age** | **Mortality rate** | **pYLD** |
| --- | --- | --- | --- | --- |
| Māori | F | 37 | 0.001121 | 0.157912 |
| Māori | F | 38 | 0.001201 | 0.157912 |
| Māori | F | 39 | 0.001311 | 0.157912 |
| Māori | F | 40 | 0.001441 | 0.160344 |
| Māori | F | 41 | 0.001611 | 0.160344 |
| Māori | F | 42 | 0.001822 | 0.160344 |
| Māori | F | 43 | 0.002062 | 0.160344 |
| Māori | F | 44 | 0.002333 | 0.160344 |
| Māori | F | 45 | 0.002643 | 0.18003 |
| Māori | F | 46 | 0.002974 | 0.18003 |
| Māori | F | 47 | 0.003346 | 0.18003 |
| Māori | F | 48 | 0.003737 | 0.18003 |
| Māori | F | 49 | 0.004149 | 0.18003 |
| Māori | F | 50 | 0.00458 | 0.17632 |
| Māori | F | 51 | 0.005033 | 0.17632 |
| Māori | F | 52 | 0.005495 | 0.17632 |
| Māori | F | 53 | 0.005998 | 0.17632 |
| Māori | F | 54 | 0.006541 | 0.17632 |
| Māori | F | 55 | 0.007156 | 0.196225 |
| Māori | F | 56 | 0.007841 | 0.196225 |
| Māori | F | 57 | 0.008617 | 0.196225 |
| Māori | F | 58 | 0.009495 | 0.196225 |
| Māori | F | 59 | 0.010495 | 0.196225 |
| Māori | F | 60 | 0.011637 | 0.228255 |
| Māori | F | 61 | 0.012923 | 0.228255 |
| Māori | F | 62 | 0.014373 | 0.228255 |
| Māori | F | 63 | 0.015977 | 0.228255 |
| Māori | F | 64 | 0.017696 | 0.228255 |
| Māori | F | 65 | 0.01954 | 0.261278 |
| Māori | F | 66 | 0.021459 | 0.261278 |
| Māori | F | 67 | 0.023463 | 0.261278 |
| Māori | F | 68 | 0.025513 | 0.261278 |
| Māori | F | 69 | 0.027608 | 0.261278 |
| Māori | F | 70 | 0.029707 | 0.284812 |
| Māori | F | 71 | 0.031811 | 0.284812 |
| Māori | F | 72 | 0.033919 | 0.284812 |
| Māori | F | 73 | 0.036125 | 0.284812 |
| Māori | F | 74 | 0.038543 | 0.284812 |
| Māori | F | 75 | 0.041312 | 0.321474 |
| Māori | F | 76 | 0.044548 | 0.321474 |
| Māori | F | 77 | 0.048371 | 0.321474 |
| Māori | F | 78 | 0.052895 | 0.321474 |
| Māori | F | 79 | 0.058254 | 0.321474 |
| Māori | F | 80 | 0.06456 | 0.375736 |
| Māori | F | 81 | 0.071947 | 0.375736 |
| Māori | F | 82 | 0.080538 | 0.375736 |
| Māori | F | 83 | 0.090373 | 0.375736 |
| Māori | F | 84 | 0.099312 | 0.375736 |
| Māori | F | 85 | 0.109614 | 0.477049 |
| Māori | F | 86 | 0.121253 | 0.477049 |
| Māori | F | 87 | 0.134206 | 0.477049 |
| Māori | F | 88 | 0.148442 | 0.477049 |
| Māori | F | 89 | 0.164038 | 0.477049 |
| Māori | F | 90 | 0.181306 | 0.477049 |
| Māori | F | 91 | 0.200661 | 0.477049 |
| Māori | F | 92 | 0.222531 | 0.477049 |
| Māori | F | 93 | 0.247347 | 0.477049 |
| Māori | F | 94 | 0.275385 | 0.477049 |
| Māori | F | 95 | 0.306023 | 0.477049 |
| Māori | F | 96 | 0.339312 | 0.477049 |
| Māori | F | 97 | 0.375319 | 0.477049 |
| Māori | F | 98 | 0.414032 | 0.477049 |
| Māori | F | 99 | 0.455407 | 0.477049 |
| Māori | F | 100 | 0.499309 | 0.477049 |
| Māori | F | 101 | 0.545572 | 0.477049 |
| Māori | F | 102 | 0.593899 | 0.477049 |
| Māori | F | 103 | 0.6439 | 0.477049 |
| Māori | F | 104 | 0.695029 | 0.477049 |
| Māori | F | 105 | 0.746632 | 0.477049 |
| Māori | F | 106 | 0.746632 | 0.477049 |
| Māori | F | 107 | 0.746632 | 0.477049 |
| Māori | F | 108 | 0.746632 | 0.477049 |
| Māori | F | 109 | 0.746632 | 0.477049 |
| Māori | F | 110 | 0.746632 | 0.477049 |

**Supplementary Table 2:** Table Initial Population Counts for MĀori Females

| Ethnicity | Sex | Age | Population |
| --- | --- | --- | --- |
| Māori | F | 0-4 | 44100 |
| Māori | F | 5-9 | 34700 |
| Māori | F | 10-14 | 33800 |
| Māori | F | 15-19 | 32600 |
| Māori | F | 20-24 | 30100 |
| Māori | F | 25-29 | 23100 |
| Māori | F | 30-34 | 21900 |
| Māori | F | 35-39 | 22300 |
| Māori | F | 40-44 | 21800 |
| Māori | F | 45-49 | 20500 |
| Māori | F | 50-54 | 17700 |
| Māori | F | 55-59 | 13100 |
| Māori | F | 60-64 | 10100 |
| Māori | F | 65-69 | 6600 |
| Māori | F | 70-74 | 5100 |
| Māori | F | 75-79 | 3100 |
| Māori | F | 80-84 | 1700 |
| Māori | F | 85-89 | 700 |
| Māori | F | 90-94 | 225 |
| Māori | F | 94-99 | 52.5 |
| Māori | F | 100-104 | 18.75 |
| Māori | F | 105-109 | 3.75 |

**Supplementary Table 3:** Mortality Rate ratios for MĀori Females

| Ethnicity | Sex | Age | Rate ratio (dep5/dep1) | Rate ratio (dep4/dep1) | Rate ratio (dep3/dep1) | Rate ratio (dep2/dep1) | Rate ratio (dep1/dep1) |
| --- | --- | --- | --- | --- | --- | --- | --- |
| Māori | F | 0-24 | 1 | 1 | 1 | 1 | 1 |
| Māori | F | 25-29 | 1.584513 | 1.438385 | 1.292256 | 1.146128 | 1 |
| Māori | F | 30-34 | 1.595621 | 1.446715 | 1.29781 | 1.148905 | 1 |
| Māori | F | 35-39 | 1.612428 | 1.459321 | 1.306214 | 1.153107 | 1 |

| Ethnicity | Sex | Age | Rate ratio<br>(dep5/dep1) | Rate ratio<br>(dep4/dep1) | Rate ratio<br>(dep3/dep1) | Rate ratio<br>(dep2/dep1) | Rate ratio<br>(dep1/dep1) |
| --- | --- | --- | --- | --- | --- | --- | --- |
| Māori | F | 40-44 | 1.635115 | 1.476336 | 1.317557 | 1.158779 | 1 |
| Māori | F | 45-49 | 1.663922 | 1.497942 | 1.331961 | 1.165981 | 1 |
| Māori | F | 50-54 | 1.699161 | 1.524371 | 1.349581 | 1.17479 | 1 |
| Māori | F | 55-59 | 1.741219 | 1.555914 | 1.37061 | 1.185305 | 1 |
| Māori | F | 60-64 | 1.581209 | 1.435907 | 1.290605 | 1.145302 | 1 |
| Māori | F | 65-69 | 1.461618 | 1.346214 | 1.230809 | 1.115405 | 1 |
| Māori | F | 70-74 | 1.343452 | 1.257589 | 1.171726 | 1.085863 | 1 |
| Māori | F | 75-79 | 1.227879 | 1.170909 | 1.113939 | 1.05697 | 1 |
| Māori | F | 80-84 | 1.115918 | 1.086938 | 1.057959 | 1.028979 | 1 |
| Māori | F | 85-89 | 1.008447 | 1.006335 | 1.004223 | 1.002112 | 1 |
| Māori | F | 90+ | 0.886413 | 0.91481 | 0.943207 | 0.971603 | 1 |

**Supplementary Table 4:** pYLD Rate ratios for MĀori Females

| Ethnicity | Sex | Age | Rate ratio<br>(dep5/dep1) | Rate ratio<br>(dep4/dep1) | Rate ratio<br>(dep3/dep1) | Rate ratio<br>(dep2/dep1) | Rate ratio<br>(dep1/dep1) |
| --- | --- | --- | --- | --- | --- | --- | --- |
| Māori | F | 0-24 | 1 | 1 | 1 | 1 | 1 |
| Māori | F | 25-29 | 1.233833 | 1.175374 | 1.116916 | 1.058458 | 1 |
| Māori | F | 30-34 | 1.187804 | 1.140853 | 1.093902 | 1.046951 | 1 |
| Māori | F | 35-39 | 1.21267 | 1.159503 | 1.106335 | 1.053168 | 1 |
| Māori | F | 40-44 | 1.175167 | 1.131375 | 1.087583 | 1.043792 | 1 |
| Māori | F | 45-49 | 1.129293 | 1.09697 | 1.064647 | 1.032323 | 1 |
| Māori | F | 50-54 | 1.157673 | 1.118255 | 1.078836 | 1.039418 | 1 |
| Māori | F | 55-59 | 1.143996 | 1.107997 | 1.071998 | 1.035999 | 1 |
| Māori | F | 60-64 | 1.127272 | 1.095454 | 1.063636 | 1.031818 | 1 |
| Māori | F | 65-69 | 1.080785 | 1.060588 | 1.040392 | 1.020196 | 1 |
| Māori | F | 70-74 | 1.064367 | 1.048275 | 1.032184 | 1.016092 | 1 |
| Māori | F | 75-79 | 1.050946 | 1.03821 | 1.025473 | 1.012737 | 1 |
| Māori | F | 80-84 | 1.042091 | 1.031568 | 1.021045 | 1.010523 | 1 |
| Māori | F | 85-89 | 1.006856 | 1.005142 | 1.003428 | 1.001714 | 1 |
| Māori | F | 90+ | 0.848308 | 0.886231 | 0.924154 | 0.962077 | 1 |

**Supplementary Table 5:** CHD rates for MĀori Females

| Ethnicity | Sex | Age | Prevalence | Incidence | Fatality | Disability Rate |
| --- | --- | --- | --- | --- | --- | --- |
| Māori | F | 0 | 0 | 0 | 0 | 0.096268 |
| Māori | F | 1 | 0 | 0 | 0 | 0.096268 |
| Māori | F | 2 | 0 | 0 | 0 | 0.096268 |
| Māori | F | 3 | 0 | 0 | 0 | 0.096268 |
| Māori | F | 4 | 0 | 0 | 0 | 0.096268 |
| Māori | F | 5 | 0 | 0 | 0 | 0.096268 |
| Māori | F | 6 | 0 | 0 | 0 | 0.096268 |
| Māori | F | 7 | 0 | 0 | 0 | 0.096268 |
| Māori | F | 8 | 0 | 0 | 0 | 0.096268 |
| Māori | F | 9 | 0 | 0 | 0 | 0.096268 |
| Māori | F | 10 | 0 | 0 | 0 | 0.096268 |
| Māori | F | 11 | 0 | 0 | 0 | 0.096268 |
| Māori | F | 12 | 0 | 0 | 0 | 0.096268 |
| Māori | F | 13 | 0 | 0 | 0.005431 | 0.096268 |
| Māori | F | 14 | 0 | 0 | 0.016293 | 0.096268 |
| Māori | F | 15 | 0 | 0 | 0.032585 | 0.096268 |
| Māori | F | 16 | 0 | 0 | 0.054309 | 0.096268 |
| Māori | F | 17 | 0 | 0 | 0.081463 | 0.096268 |
| Māori | F | 18 | 0 | 0 | 0.114048 | 0.096268 |
| Māori | F | 19 | 8.6531E-05 | 4.65E-05 | 0.142915 | 0.096268 |
| Māori | F | 20 | 0.00012766 | 6.9E-05 | 0.162633 | 0.096268 |
| Māori | F | 21 | 0.00018458 | 9.78E-05 | 0.173202 | 0.096268 |
| Māori | F | 22 | 0.0002609 | 0.000133 | 0.174623 | 0.096268 |
| Māori | F | 23 | 0.0003611 | 0.000174 | 0.166894 | 0.096268 |
| Māori | F | 24 | 0.00049148 | 0.000221 | 0.150016 | 0.096268 |
| Māori | F | 25 | 0.00065551 | 0.000264 | 0.126389 | 0.096268 |
| Māori | F | 26 | 0.00084868 | 0.000295 | 0.105162 | 0.096268 |
| Māori | F | 27 | 0.00106311 | 0.000315 | 0.086334 | 0.096268 |
| Māori | F | 28 | 0.00129157 | 0.000324 | 0.069907 | 0.096268 |
| Māori | F | 29 | 0.00152665 | 0.000322 | 0.055878 | 0.096268 |
| Māori | F | 30 | 0.00176014 | 0.000309 | 0.04425 | 0.096268 |
| Māori | F | 31 | 0.00198558 | 0.000294 | 0.036755 | 0.096268 |
| Māori | F | 32 | 0.00220548 | 0.000288 | 0.030996 | 0.096268 |
| Māori | F | 33 | 0.0024283 | 0.000292 | 0.02697 | 0.096268 |
| Māori | F | 34 | 0.00266111 | 0.000306 | 0.024679 | 0.096268 |
| Māori | F | 35 | 0.00290969 | 0.000329 | 0.024122 | 0.096268 |
| Māori | F | 36 | 0.00317849 | 0.000361 | 0.0253 | 0.096268 |
| Māori | F | 37 | 0.00348683 | 0.000434 | 0.027466 | 0.096268 |
| Māori | F | 38 | 0.00386711 | 0.000537 | 0.028885 | 0.096268 |
| Māori | F | 39 | 0.00434901 | 0.000671 | 0.029557 | 0.096268 |
| Māori | F | 40 | 0.004962 | 0.000836 | 0.029484 | 0.096268 |
| Māori | F | 41 | 0.00573568 | 0.001031 | 0.028663 | 0.096268 |
| Māori | F | 42 | 0.00670044 | 0.001257 | 0.027096 | 0.096268 |
| Māori | F | 43 | 0.00789852 | 0.001537 | 0.02504 | 0.096268 |
| Māori | F | 44 | 0.00936581 | 0.001839 | 0.02324 | 0.096268 |
| Māori | F | 45 | 0.01112029 | 0.002165 | 0.021698 | 0.11249 |
| Māori | F | 46 | 0.01317935 | 0.002514 | 0.020412 | 0.11249 |
| Māori | F | 47 | 0.01555942 | 0.002887 | 0.019383 | 0.11249 |
| Māori | F | 48 | 0.01827551 | 0.003282 | 0.018612 | 0.11249 |
| Māori | F | 49 | 0.02132469 | 0.003667 | 0.018112 | 0.11249 |
| Māori | F | 50 | 0.02467794 | 0.00402 | 0.017628 | 0.11249 |
| Māori | F | 51 | 0.02829785 | 0.004339 | 0.01716 | 0.11249 |
| Māori | F | 52 | 0.03214814 | 0.004625 | 0.016707 | 0.11249 |
| Māori | F | 53 | 0.0361936 | 0.004879 | 0.016269 | 0.11249 |
| Māori | F | 54 | 0.04040007 | 0.005099 | 0.015847 | 0.11249 |
| Māori | F | 55 | 0.04478414 | 0.005407 | 0.015777 | 0.123975 |
| Māori | F | 56 | 0.04942521 | 0.005835 | 0.016043 | 0.123975 |
| Māori | F | 57 | 0.05441109 | 0.006384 | 0.016646 | 0.123975 |
| Māori | F | 58 | 0.05982106 | 0.007053 | 0.017586 | 0.123975 |
| Māori | F | 59 | 0.06572488 | 0.007842 | 0.018862 | 0.123975 |
| Māori | F | 60 | 0.07218178 | 0.008751 | 0.020475 | 0.123975 |
| Māori | F | 61 | 0.07921749 | 0.009712 | 0.02216 | 0.123975 |
| Māori | F | 62 | 0.08678621 | 0.010604 | 0.023583 | 0.123975 |
| Māori | F | 63 | 0.09480191 | 0.011427 | 0.024743 | 0.123975 |
| Māori | F | 64 | 0.10318604 | 0.012181 | 0.025639 | 0.123975 |
| Māori | F | 65 | 0.11186833 | 0.012867 | 0.026272 | 0.109541 |
| Māori | F | 66 | 0.12078727 | 0.013483 | 0.026642 | 0.109541 |
| Māori | F | 67 | 0.12988245 | 0.01403 | 0.026881 | 0.109541 |
| Māori | F | 68 | 0.13910704 | 0.014576 | 0.027255 | 0.109541 |
| Māori | F | 69 | 0.14843047 | 0.015121 | 0.027761 | 0.109541 |
| Māori | F | 70 | 0.15782072 | 0.015666 | 0.028402 | 0.109541 |
| Māori | F | 71 | 0.1672445 | 0.01621 | 0.029175 | 0.109541 |
| Māori | F | 72 | 0.17666751 | 0.016753 | 0.030082 | 0.109541 |
| Māori | F | 73 | 0.18608633 | 0.017485 | 0.031713 | 0.109541 |
| Māori | F | 74 | 0.19553112 | 0.018407 | 0.033934 | 0.109541 |
| Māori | F | 75 | 0.2050256 | 0.01952 | 0.036745 | 0.100756 |
| Māori | F | 76 | 0.21457837 | 0.020823 | 0.040147 | 0.100756 |
| Māori | F | 77 | 0.22418386 | 0.022316 | 0.04414 | 0.100756 |
| Māori | F | 78 | 0.23382357 | 0.023999 | 0.048723 | 0.100756 |
| Māori | F | 79 | 0.24337891 | 0.025619 | 0.053848 | 0.100756 |
| Māori | F | 80 | 0.25262657 | 0.026986 | 0.058927 | 0.100756 |
| Māori | F | 81 | 0.26134547 | 0.028099 | 0.063958 | 0.100756 |
| Māori | F | 82 | 0.26934014 | 0.028959 | 0.068941 | 0.100756 |
| Māori | F | 83 | 0.27643974 | 0.029566 | 0.073878 | 0.100756 |
| Māori | F | 84 | 0.28249618 | 0.029919 | 0.078766 | 0.100756 |
| Māori | F | 85 | 0.28723392 | 0.029987 | 0.085003 | 0.089604 |
| Māori | F | 86 | 0.29033412 | 0.030022 | 0.092634 | 0.089604 |
| Māori | F | 87 | 0.29158889 | 0.030024 | 0.10166 | 0.089604 |
| Māori | F | 88 | 0.29081797 | 0.029995 | 0.112082 | 0.089604 |
| Māori | F | 89 | 0.28787713 | 0.029932 | 0.123898 | 0.089604 |
| Māori | F | 90 | 0.28266789 | 0.029838 | 0.137109 | 0.089604 |
| Māori | F | 91 | 0.27504134 | 0.030028 | 0.154038 | 0.089604 |
| Māori | F | 92 | 0.26497195 | 0.030535 | 0.17329 | 0.089604 |
| Māori | F | 93 | 0.25270519 | 0.03136 | 0.194863 | 0.089604 |
| Māori | F | 94 | 0.23860577 | 0.032503 | 0.21876 | 0.089604 |
| Māori | F | 95 | 0.2231577 | 0.033962 | 0.244978 | 0.089604 |
| Māori | F | 96 | 0.20694661 | 0.035739 | 0.27352 | 0.089604 |
| Māori | F | 97 | 0.19080747 | 0.037535 | 0.299974 | 0.089604 |
| Māori | F | 98 | 0.17544101 | 0.039509 | 0.329074 | 0.089604 |
| Māori | F | 99 | 0.16288367 | 0.041051 | 0.35599 | 0.089604 |
| Māori | F | 100 | 0.16288367 | 0.041051 | 0.35599 | 0.089604 |
| Māori | F | 101 | 0.16288367 | 0.041051 | 0.35599 | 0.089604 |
| Māori | F | 102 | 0.16288367 | 0.041051 | 0.35599 | 0.089604 |
| Māori | F | 103 | 0.16288367 | 0.041051 | 0.35599 | 0.089604 |
| Māori | F | 104 | 0.16288367 | 0.041051 | 0.35599 | 0.089604 |
| Māori | F | 105 | 0.16288367 | 0.041051 | 0.35599 | 0.089604 |
| Māori | F | 106 | 0.16288367 | 0.041051 | 0.35599 | 0.089604 |
| Māori | F | 107 | 0.16288367 | 0.041051 | 0.35599 | 0.089604 |
| Māori | F | 108 | 0.16288367 | 0.041051 | 0.35599 | 0.089604 |
| Māori | F | 109 | 0.16288367 | 0.041051 | 0.35599 | 0.089604 |
| Māori | F | 110 | 0.16288367 | 0.041051 | 0.35599 | 0.089604 |

**Supplementary Table 6:** CHD Incidence rate ratios for MĀori Females

| Ethnicity | Sex | Age | Rate ratio<br>(dep5/dep1) | Rate ratio<br>(dep4/dep1) | Rate ratio<br>(dep3/dep1) | Rate ratio<br>(dep2/dep1) | Rate ratio<br>(dep1/dep1) |
| --- | --- | --- | --- | --- | --- | --- | --- |
| Māori | F | 0-24 | 1 | 1 | 1 | 1 | 1 |
| Māori | F | 25-29 | 2.508118 | 2.131088 | 1.754059 | 1.377029 | 1 |
| Māori | F | 30-34 | 2.447497 | 2.085623 | 1.723748 | 1.361874 | 1 |
| Māori | F | 35-39 | 2.359299 | 2.019474 | 1.67965 | 1.339825 | 1 |
| Māori | F | 40-44 | 2.246627 | 1.93497 | 1.623314 | 1.311657 | 1 |
| Māori | F | 45-49 | 2.113324 | 1.834993 | 1.556662 | 1.278331 | 1 |
| Māori | F | 50-54 | 1.963758 | 1.722819 | 1.481879 | 1.24094 | 1 |
| Māori | F | 55-59 | 1.80259 | 1.601942 | 1.401295 | 1.200647 | 1 |
| Māori | F | 60-64 | 1.717948 | 1.538461 | 1.358974 | 1.179487 | 1 |
| Māori | F | 65-69 | 1.533512 | 1.400134 | 1.266756 | 1.133378 | 1 |
| Māori | F | 70-74 | 1.384246 | 1.288185 | 1.192123 | 1.096062 | 1 |
| Māori | F | 75-79 | 1.26354 | 1.197655 | 1.13177 | 1.065885 | 1 |
| Māori | F | 80-84 | 1.166309 | 1.124732 | 1.083154 | 1.041577 | 1 |
| Māori | F | 85-89 | 1.088648 | 1.066486 | 1.044324 | 1.022162 | 1 |
| Māori | F | 90+ | 1.017131 | 1.012848 | 1.008565 | 1.004283 | 1 |

### Disaggregation input parameters

The data in supplementary tables 6, 8, 10, and 12 were based on extracts from the following NZ national health collections: National Health Index (NHI), Primary Health Organisation (PHO) Enrolment collection, Laboratory Claims, Pharmaceutical Claims, National Minimum Dataset (NMDS) - publicly funded hospitalizations only, Cancer Registry, Mortality Collection, National Non-Admitted Patient Collection, General Medical Subsidy (GMS) Claims.

#### Health Service user population, and disease indications

We created a health service user population for the 2011/12 financial year based on recorded contact and residence status in the above collections. Amongst this group of people, we used the algorithms described below to flag whether or not an individual had shown an indication of CHD or stroke in 2011/12:

##### CHD

People with a publicly-funded hospital discharge with any (primary or otherwise) diagnosis of ICD-10-AM codes I20-I25 on or before the specified financial year who were alive for at least part of the year, or with an underlying cause of death recorded as one of these ICD-10-codes within the specified financial year, or with one of the following publicly-funded hospital procedures during the specified financial year:

(ICD-10-AM: 3530400, 3530500, 3531000, 3531001, 3531002, 3849700, 3849701, 3849702, 3849703, 3849704, 3849705, 3849706, 3849707, 3850000, 3850001, 3850002, 3850003, 3850004, 3850300, 3850301, 3850302, 3850303, 3850304, 3863700, 9020100, 9020101, 9020102, 9020103)

or with two or more dispensings of one of the following pharmaceuticals :

(Glyceryl trinitrate 1577, Isosorbide Dinitrate 2377, Isosorbide mononitrate 2836, Nicorandil 1272, Perhexiline maleate 1949) in the specified financial year

##### Stroke

People with a publicly-funded hospital discharge with any (primary or otherwise) diagnosis of ICD-10-AM codes G45-G46, I60-I67 on or before the specified financial year who were alive for at least part of the year, or with an underlying cause of death recorded as one of these ICD-10-codes, during the specified financial year

#### NZ Deprivation quintile

We created a variable to flag NZ Deprivation Index quintile (as there is no standard NZDep variable as part of these collections) based on the following method:

We extracted all domicile codes for each person for the years 2004/05 to 2014/15, along with relevant dates, from the following collections: NMDS (event end date), NHI (last updated date), Cancer registrations (diagnosis date), Mortality (date of death), NNPAC (date of service), PHO (last consultation date). Note that one person can have many domicile codes over a year.

The domicile codes were mapped to census area units, which in turn were mapped to NZ Dep quintiles, first using 2013 mappings (and then if they didn’t work, 2006 mappings). Where an individual in the health service user population had more than one NZDep quintile value for a particular financial year, we assigned the quintile value closest to the end of the financial year. Where an individual did not have an NZDep quintile for a particular financial year, we used an NZDep quintile from the closest financial year (looking back first). A maximum five year range was used (that is, two years either side of the particular year). Finally, the NZDep quintile based on the domicile code from the NHI – which represented the NZDep from the last recorded (i.e. most recent) domicile code was added on to fill any remaining gaps.

#### Incidence

Incidence was measured as counts of people with their first indication of CHD or Stroke in 2011/12 divided by person time at risk (based on the health service use population). When incidence was calculated by age, sex, ethnic, and deprivation quintile group, the numbers were very small and patterns in rates were unstable. Predicted rates were calculated using Poisson regression (or negative binomial if overdispersed).

#### Prevalence

Prevalence was calculated as a proportion with number of cases (of CHD or stroke) divided by the health service user population. As for the incidence measure, rates were unstable when broken down by the four demographic variables, so logistic regression was used to calculate predicted rates.

**Supplementary Table 7:** CHD Fatality rate ratios for MĀori Females

| Ethnicity | Sex | Age | Rate ratio<br>(dep5/dep1) | Rate ratio<br>(dep4/dep1) | Rate ratio<br>(dep3/dep1) | Rate ratio<br>(dep2/dep1) | Rate ratio<br>(dep1/dep1) |
| --- | --- | --- | --- | --- | --- | --- | --- |
| Māori | F | 0-24 | 1 | 1 | 1 | 1 | 1 |
| Māori | F | 25-29 | 1.48253 | 1.361897 | 1.241265 | 1.120632 | 1 |
| Māori | F | 30-34 | 1.456976 | 1.342732 | 1.228488 | 1.114244 | 1 |
| Māori | F | 35-39 | 1.419471 | 1.314603 | 1.209735 | 1.104868 | 1 |
| Māori | F | 40-44 | 1.370961 | 1.27822 | 1.18548 | 1.09274 | 1 |
| Māori | F | 45-49 | 1.312648 | 1.984486 | 1.156324 | 1.078162 | 1 |
| Māori | F | 50-54 | 1.245937 | 1.934453 | 1.122969 | 1.061484 | 1 |
| Māori | F | 55-59 | 1.172381 | 1.879286 | 1.086191 | 1.043095 | 1 |
| Māori | F | 60-64 | 1.187543 | 1.890657 | 1.093771 | 1.046886 | 1 |
| Māori | F | 65-69 | 1.189493 | 1.892119 | 1.094746 | 1.047373 | 1 |
| Māori | F | 70-74 | 1.167084 | 1.125313 | 1.083542 | 1.041771 | 1 |
| Māori | F | 75-79 | 1.121684 | 1.091263 | 1.060842 | 1.030421 | 1 |
| Māori | F | 80-84 | 1.056009 | 1.042007 | 1.028005 | 1.014002 | 1 |
| Māori | F | 85-89 | 0.973852 | 0.980389 | 0.986926 | 0.993463 | 1 |
| Māori | F | 90+ | 0.859885 | 0.894913 | 0.929942 | 0.964971 | 1 |

**Supplementary Table 8:** CHD Prevalence ratios for MĀori Females

| Ethnicity | Sex | Age | Rate ratio<br>(dep5/dep1) | Rate ratio<br>(dep4/dep1) | Rate ratio<br>(dep3/dep1) | Rate ratio<br>(dep2/dep1) | Rate ratio<br>(dep1/dep1) |
| --- | --- | --- | --- | --- | --- | --- | --- |
| Māori | F | 0-24 | 1 | 1 | 1 | 1 | 1 |
| Māori | F | 25-29 | 2.137763 | 1.853322 | 1.568881 | 1.284441 | 1 |
| Māori | F | 30-34 | 2.127309 | 1.845482 | 1.563655 | 1.281827 | 1 |
| Māori | F | 35-39 | 2.102494 | 1.826871 | 1.551247 | 1.275624 | 1 |
| Māori | F | 40-44 | 2.063819 | 1.797864 | 1.53191 | 1.265955 | 1 |
| Māori | F | 45-49 | 2.012061 | 1.759046 | 1.50603 | 1.253015 | 1 |
| Māori | F | 50-54 | 1.948246 | 1.711185 | 1.474123 | 1.237062 | 1 |
| Māori | F | 55-59 | 1.873609 | 1.655207 | 1.436805 | 1.218402 | 1 |
| Māori | F | 60-64 | 1.697884 | 1.523413 | 1.348942 | 1.174471 | 1 |
| Māori | F | 65-69 | 1.483933 | 1.36295 | 1.241967 | 1.120983 | 1 |
| Māori | F | 70-74 | 1.323865 | 1.242899 | 1.161933 | 1.080966 | 1 |
| Māori | F | 75-79 | 1.205583 | 1.154187 | 1.102791 | 1.051396 | 1 |
| Māori | F | 80-84 | 1.120657 | 1.090493 | 1.060329 | 1.030164 | 1 |
| Māori | F | 85-89 | 1.063342 | 1.047506 | 1.031671 | 1.015835 | 1 |
| Māori | F | 90+ | 1.025867 | 1.019401 | 1.012934 | 1.006467 | 1 |

**Supplementary Table 9:** Stroke rates for MĀori Females

| Ethnicity | Sex | Age | Prevalence | Incidence | Fatality | Disability Rate |
| --- | --- | --- | --- | --- | --- | --- |
| Māori | F | 0 | 0 | 0 | 0 | 0.431037 |
| Māori | F | 1 | 0 | 0 | 0 | 0.431037 |
| Māori | F | 2 | 0 | 0 | 0 | 0.431037 |
| Māori | F | 3 | 0 | 0 | 0 | 0.431037 |
| Māori | F | 4 | 0 | 0 | 0 | 0.431037 |
| Māori | F | 5 | 0 | 0 | 0 | 0.431037 |
| Māori | F | 6 | 0 | 0 | 0 | 0.431037 |
| Māori | F | 7 | 0 | 0 | 0 | 0.431037 |
| Māori | F | 8 | 0 | 0 | 0 | 0.431037 |
| Māori | F | 9 | 0 | 0 | 0 | 0.431037 |
| Māori | F | 10 | 0 | 0 | 0 | 0.431037 |
| Māori | F | 11 | 0 | 0 | 0 | 0.431037 |
| Māori | F | 12 | 0 | 0 | 0 | 0.431037 |
| Māori | F | 13 | 0 | 0 | 0 | 0.431037 |
| Māori | F | 14 | 0 | 0 | 0 | 0.431037 |
| Māori | F | 15 | 0 | 0 | 0 | 0.431037 |
| Māori | F | 16 | 0 | 0 | 0 | 0.431037 |
| Māori | F | 17 | 0 | 0 | 0 | 0.431037 |
| Māori | F | 18 | 0 | 0 | 0 | 0.431037 |
| Māori | F | 19 | 0 | 0 | 0 | 0.431037 |
| Māori | F | 20 | 0 | 0 | 0 | 0.431037 |
| Māori | F | 21 | 0 | 0 | 0 | 0.431037 |
| Māori | F | 22 | 0 | 0 | 0 | 0.431037 |
| Māori | F | 23 | 0 | 0 | 0 | 0.431037 |
| Māori | F | 24 | 0 | 0 | 0 | 0.431037 |
| Māori | F | 25 | 5.48E-07 | 1.12E-06 | 0.047619 | 0.431037 |
| Māori | F | 26 | 2.59E-06 | 3.37E-06 | 0.142857 | 0.431037 |
| Māori | F | 27 | 6.5E-06 | 6.73E-06 | 0.285714 | 0.431037 |
| Māori | F | 28 | 1.17E-05 | 1.12E-05 | 0.47619 | 0.431037 |
| Māori | F | 29 | 1.68E-05 | 1.68E-05 | 0.714286 | 0.431037 |
| Māori | F | 30 | 2.05E-05 | 2.36E-05 | 1 | 0.431037 |
| Māori | F | 31 | 2.3E-05 | 3.2E-05 | 1.285714 | 0.431037 |
| Māori | F | 32 | 2.52E-05 | 4.1E-05 | 1.52381 | 0.431037 |
| Māori | F | 33 | 2.76E-05 | 5.07E-05 | 1.714286 | 0.431037 |
| Māori | F | 34 | 3.06E-05 | 6.09E-05 | 1.857143 | 0.431037 |
| Māori | F | 35 | 3.41E-05 | 7.17E-05 | 1.952381 | 0.431037 |
| Māori | F | 36 | 3.84E-05 | 8.31E-05 | 2 | 0.431037 |
| Māori | F | 37 | 5.03E-05 | 0.000123 | 1.95321 | 0.431037 |
| Māori | F | 38 | 7.8E-05 | 0.000191 | 1.859631 | 0.431037 |
| Māori | F | 39 | 0.000125 | 0.000287 | 1.719261 | 0.431037 |
| Māori | F | 40 | 0.000199 | 0.000411 | 1.532102 | 0.431037 |
| Māori | F | 41 | 0.000313 | 0.000564 | 1.298153 | 0.431037 |
| Māori | F | 42 | 0.000493 | 0.000744 | 1.017414 | 0.431037 |
| Māori | F | 43 | 0.000781 | 0.000943 | 0.736539 | 0.431037 |
| Māori | F | 44 | 0.001213 | 0.001133 | 0.502318 | 0.431037 |
| Māori | F | 45 | 0.001834 | 0.001315 | 0.314749 | 0.137442 |
| Māori | F | 46 | 0.002705 | 0.001487 | 0.173835 | 0.137442 |
| Māori | F | 47 | 0.003879 | 0.001651 | 0.079573 | 0.137442 |
| Māori | F | 48 | 0.005359 | 0.001805 | 0.031965 | 0.137442 |
| Māori | F | 49 | 0.007026 | 0.001939 | 0.031062 | 0.137442 |
| Māori | F | 50 | 0.00877 | 0.002061 | 0.030209 | 0.137442 |
| Māori | F | 51 | 0.010581 | 0.002172 | 0.029406 | 0.137442 |
| Māori | F | 52 | 0.012447 | 0.002272 | 0.028654 | 0.137442 |
| Māori | F | 53 | 0.014358 | 0.00236 | 0.027953 | 0.137442 |
| Māori | F | 54 | 0.016302 | 0.002436 | 0.027303 | 0.137442 |
| Māori | F | 55 | 0.018276 | 0.002517 | 0.026902 | 0.129704 |
| Māori | F | 56 | 0.020284 | 0.002613 | 0.0267 | 0.129704 |
| Māori | F | 57 | 0.022339 | 0.002725 | 0.026697 | 0.129704 |
| Māori | F | 58 | 0.02445 | 0.002853 | 0.026893 | 0.129704 |
| Māori | F | 59 | 0.026625 | 0.002996 | 0.027287 | 0.129704 |
| Māori | F | 60 | 0.028869 | 0.003155 | 0.02788 | 0.129704 |
| Māori | F | 61 | 0.031193 | 0.003338 | 0.02869 | 0.129704 |
| Māori | F | 62 | 0.033602 | 0.00353 | 0.029518 | 0.129704 |
| Māori | F | 63 | 0.036098 | 0.003731 | 0.030364 | 0.129704 |
| Māori | F | 64 | 0.03868 | 0.003941 | 0.031228 | 0.129704 |
| Māori | F | 65 | 0.04135 | 0.004159 | 0.03211 | 0.163902 |
| Māori | F | 66 | 0.044107 | 0.004387 | 0.03301 | 0.163902 |
| Māori | F | 67 | 0.046947 | 0.004626 | 0.034107 | 0.163902 |
| Māori | F | 68 | 0.049861 | 0.004867 | 0.035384 | 0.163902 |
| Māori | F | 69 | 0.052832 | 0.005111 | 0.036841 | 0.163902 |
| Māori | F | 70 | 0.055843 | 0.005357 | 0.038478 | 0.163902 |
| Māori | F | 71 | 0.058878 | 0.005606 | 0.040295 | 0.163902 |
| Māori | F | 72 | 0.061917 | 0.005858 | 0.042291 | 0.163902 |
| Māori | F | 73 | 0.064941 | 0.006115 | 0.044522 | 0.163902 |
| Māori | F | 74 | 0.067936 | 0.006376 | 0.046807 | 0.163902 |
| Māori | F | 75 | 0.070889 | 0.00664 | 0.049147 | 0.258457 |
| Māori | F | 76 | 0.07379 | 0.006908 | 0.051542 | 0.258457 |
| Māori | F | 77 | 0.076629 | 0.007179 | 0.053991 | 0.258457 |
| Māori | F | 78 | 0.079396 | 0.007453 | 0.056495 | 0.258457 |
| Māori | F | 79 | 0.082061 | 0.007898 | 0.061678 | 0.258457 |
| Māori | F | 80 | 0.084567 | 0.008512 | 0.069485 | 0.258457 |
| Māori | F | 81 | 0.086845 | 0.009293 | 0.079916 | 0.258457 |
| Māori | F | 82 | 0.088821 | 0.010243 | 0.092971 | 0.258457 |
| Māori | F | 83 | 0.090421 | 0.01136 | 0.10865 | 0.258457 |
| Māori | F | 84 | 0.091581 | 0.012646 | 0.126953 | 0.258457 |
| Māori | F | 85 | 0.092002 | 0.014028 | 0.153503 | 0.558867 |
| Māori | F | 86 | 0.091237 | 0.015338 | 0.185677 | 0.558867 |
| Māori | F | 87 | 0.089015 | 0.016576 | 0.223474 | 0.558867 |
| Māori | F | 88 | 0.085244 | 0.017742 | 0.266895 | 0.558867 |
| Māori | F | 89 | 0.080037 | 0.018837 | 0.315939 | 0.558867 |
| Māori | F | 90 | 0.073694 | 0.019861 | 0.370606 | 0.558867 |
| Māori | F | 91 | 0.066711 | 0.020792 | 0.428389 | 0.558867 |
| Māori | F | 92 | 0.059802 | 0.021703 | 0.483665 | 0.558867 |
| Māori | F | 93 | 0.053618 | 0.022593 | 0.536433 | 0.558867 |
| Māori | F | 94 | 0.048464 | 0.023463 | 0.586694 | 0.558867 |
| Māori | F | 95 | 0.044383 | 0.024313 | 0.634447 | 0.558867 |
| Māori | F | 96 | 0.041262 | 0.025143 | 0.679693 | 0.558867 |
| Māori | F | 97 | 0.038963 | 0.025836 | 0.716325 | 0.558867 |
| Māori | F | 98 | 0.037271 | 0.026599 | 0.756621 | 0.558867 |
| Māori | F | 99 | 0.036039 | 0.02712 | 0.793784 | 0.558867 |
| Māori | F | 100 | 0.036039 | 0.02712 | 0.793784 | 0.558867 |
| Māori | F | 101 | 0.036039 | 0.02712 | 0.793784 | 0.558867 |
| Māori | F | 102 | 0.036039 | 0.02712 | 0.793784 | 0.558867 |
| Māori | F | 103 | 0.036039 | 0.02712 | 0.793784 | 0.558867 |
| Māori | F | 104 | 0.036039 | 0.02712 | 0.793784 | 0.558867 |
| Māori | F | 105 | 0.036039 | 0.02712 | 0.793784 | 0.558867 |
| Māori | F | 106 | 0.036039 | 0.02712 | 0.793784 | 0.558867 |
| Māori | F | 107 | 0.036039 | 0.02712 | 0.793784 | 0.558867 |
| Māori | F | 108 | 0.036039 | 0.02712 | 0.793784 | 0.558867 |
| Māori | F | 109 | 0.036039 | 0.02712 | 0.793784 | 0.558867 |
| Māori | F | 110 | 0.036039 | 0.02712 | 0.793784 | 0.558867 |

**Supplementary Table 10:** Stroke Incidence rate ratios for MĀori Females

| Ethnicity | Sex | Age | Rate ratio (dep5/dep1) | Rate ratio (dep4/dep1) | Rate ratio (dep3/dep1) | Rate ratio (dep2/dep1) | Rate ratio (dep1/dep1) |
| --- | --- | --- | --- | --- | --- | --- | --- |
| Māori | F | 0-24 | 1 | 1 | 1 | 1 | 1 |
| Māori | F | 25-29 | 1.573997 | 1.430498 | 1.286999 | 1.143499 | 1 |
| Māori | F | 30-34 | 1.597383 | 1.448037 | 1.298691 | 1.149346 | 1 |
| Māori | F | 35-39 | 1.633113 | 1.474835 | 1.316557 | 1.158278 | 1 |
| Māori | F | 40-44 | 1.682 | 1.5115 | 1.341 | 1.1705 | 1 |
| Māori | F | 45-49 | 1.745172 | 1.558879 | 1.372586 | 1.186293 | 1 |
| Māori | F | 50-54 | 1.824119 | 1.618089 | 1.412059 | 1.20603 | 1 |
| Māori | F | 55-59 | 1.920749 | 1.690562 | 1.460375 | 1.230187 | 1 |
| Māori | F | 60-64 | 1.663498 | 1.497623 | 1.331749 | 1.165874 | 1 |
| Māori | F | 65-69 | 1.438686 | 1.329014 | 1.219343 | 1.109671 | 1 |
| Māori | F | 70-74 | 1.273003 | 1.204752 | 1.136501 | 1.068251 | 1 |
| Māori | F | 75-79 | 1.152425 | 1.114319 | 1.076212 | 1.038106 | 1 |
| Māori | F | 80-84 | 1.067372 | 1.050529 | 1.033686 | 1.016843 | 1 |
| Māori | F | 85-89 | 1.011437 | 1.008578 | 1.005719 | 1.002859 | 1 |
| Māori | F | 90+ | 0.977196 | 0.982897 | 0.988598 | 0.994299 | 1 |

**Supplementary Table 11:**
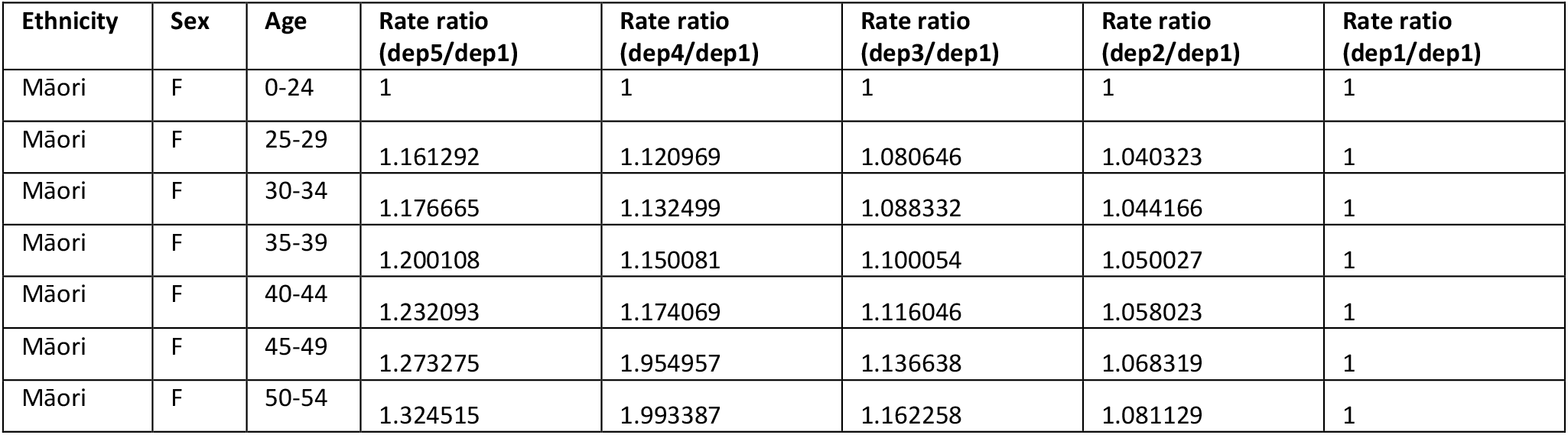
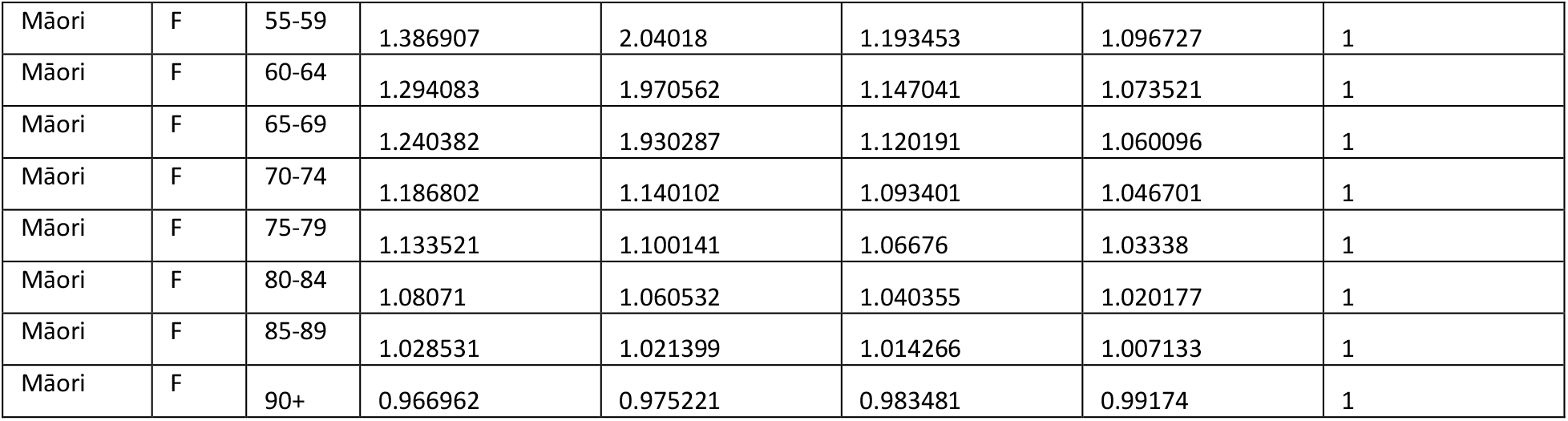
Stroke Fatality rate ratios for MĀori Females

**Supplementary Table 12:** Stroke Prevalence ratios for MĀori Females

| Ethnicity | Sex | Age | Rate ratio (dep5/dep1) | Rate ratio (dep4/dep1) | Rate ratio (dep3/dep1) | Rate ratio (dep2/dep1) | Rate ratio (dep1/dep1) |
| --- | --- | --- | --- | --- | --- | --- | --- |
| Māori | F | 0-24 | 1 | 1 | 1 | 1 | 1 |
| Māori | F | 25-29 | 1.549719 | 1.412289 | 1.27486 | 1.13743 | 1 |
| Māori | F | 30-34 | 1.58028 | 1.43521 | 1.29014 | 1.14507 | 1 |
| Māori | F | 35-39 | 1.621962 | 1.466471 | 1.310981 | 1.15549 | 1 |
| Māori | F | 40-44 | 1.675606 | 1.506704 | 1.337803 | 1.168901 | 1 |
| Māori | F | 45-49 | 1.742323 | 1.556742 | 1.371161 | 1.185581 | 1 |
| Māori | F | 50-54 | 1.823519 | 1.617639 | 1.411759 | 1.20588 | 1 |
| Māori | F | 55-59 | 1.920954 | 1.690716 | 1.460477 | 1.230239 | 1 |
| Māori | F | 60-64 | 1.660236 | 1.495177 | 1.330118 | 1.165059 | 1 |
| Māori | F | 65-69 | 1.428493 | 1.321369 | 1.214246 | 1.107123 | 1 |
| Māori | F | 70-74 | 1.261287 | 1.195966 | 1.130644 | 1.065322 | 1 |
| Māori | F | 75-79 | 1.142819 | 1.107114 | 1.07141 | 1.035705 | 1 |
| Māori | F | 80-84 | 1.062598 | 1.046948 | 1.031299 | 1.015649 | 1 |
| Māori | F | 85-89 | 1.013883 | 1.010412 | 1.006941 | 1.003471 | 1 |
| Māori | F | 90+ | 0.991632 | 0.993724 | 0.995816 | 0.997908 | 1 |

**Supplementary Table 13:** Sodium Daily Intake Proportions for MĀori Females (rows sum to 1)

| Ethnicity | Sex | Age | 0-1 gram | 1-2 gram | 2-3 gram | 3-4 gram | 4-5 gram | 5-6 gram | 6-7 gram | >7 gram |
| --- | --- | --- | --- | --- | --- | --- | --- | --- | --- | --- |
| Māori | F | 0-24 | 0.028985507 | 0.217391304 | 0.297101449 | 0.188405797 | 0.115942029 | 0.072463768 | 0.036231884 | 0.043478261 |
| Māori | F | 25-34 | 0.030303 | 0.187879 | 0.218182 | 0.218182 | 0.175758 | 0.090909 | 0.030303 | 0.048485 |
| Māori | F | 35-44 | 0.02963 | 0.2 | 0.266667 | 0.2 | 0.125926 | 0.111111 | 0.02963 | 0.037037 |
| Māori | F | 45-54 | 0.08642 | 0.160494 | 0.333333 | 0.17284 | 0.111111 | 0.049383 | 0.074074 | 0.012346 |
| Māori | F | 55-64 | 0.033333 | 0.283333 | 0.266667 | 0.216667 | 0.1 | 0.05 | 0.016667 | 0.033333 |
| Māori | F | 65-74 | 0.051282 | 0.333333 | 0.333333 | 0.179487 | 0.025641 | 0.051282 | 0.025641 | 0 |
| Māori | F | 75-84 | 0.071429 | 0.428571 | 0.214286 | 0.071429 | 0.071429 | 0.071429 | 0 | 0.071429 |
| Māori | F | 85+ | 0 | 0 | 0.333333 | 0.666667 | 0 | 0 | 0 | 0 |

**Supplementary Table 14:** Relative Risks for CHD and Stroke per 1g/day Sodium Increase for MĀori Females*

| Ethnicity | Sex | Age | CHD | Stroke |
| --- | --- | --- | --- | --- |
| Māori | F | 0-24 | 1 | 1 |
| Māori | F | 25-29 | 1.04 | 1.05182 |
| Māori | F | 30-34 | 1.05 | 1.06965 |
| Māori | F | 35-39 | 1.057 | 1.08688 |
| Māori | F | 40-44 | 1.063 | 1.10034 |
| Māori | F | 45-49 | 1.073 | 1.10901 |
| Māori | F | 50-54 | 1.082 | 1.11714 |
| Māori | F | 55-59 | 1.088 | 1.12139 |
| Māori | F | 60-64 | 1.091 | 1.12213 |
| Māori | F | 65-69 | 1.093 | 1.11908 |
| Māori | F | 70-74 | 1.084 | 1.10173 |
| Māori | F | 75-79 | 1.074 | 1.08226 |
| Māori | F | 80-84 | 1.058 | 1.04165 |
| Māori | F | 85-89 | 1.058 | 1.04156 |
| Māori | F | 90+ | 1.058 | 1.04152 |

\*With a theoretical minimum risk exposure level (TMREL) of 3g sodium/day

**Supplementary Table 15:** Sodium Daily Intake BAU Mean Exposures for MĀori Females

| Ethnicity | Sex | 0-1 gram | 1-2 gram | 2-3 gram | 3-4 gram | 4-5 gram | 5-6 gram | 6-7 gram | >7 gram |
| --- | --- | --- | --- | --- | --- | --- | --- | --- | --- |
| Māori | F | 0.678652 | 1.574601 | 2.511774 | 3.474541 | 4.441067 | 5.469723 | 6.477527 | 8.845344 |

